# Deep Learning Cerebellar Magnetic Resonance Imaging Segmentation in Late-Onset GM2 Gangliosidosis: Implications for Phenotype

**DOI:** 10.1101/2025.04.08.25325262

**Authors:** Connor J. Lewis, Selby I. Chipman, Jean M. Johnston, Maria T. Acosta, Cynthia J. Tifft, Camilo Toro

**Author notes:** Corresponding Author: Camilo Toro.

## Abstract

Late-onset Tay-Sachs (LOTS) disease and late-onset Sandhoff disease (LOSD) have long been considered indistinguishable due to similar clinical presentations and shared biochemical deficits. However, recent magnetic resonance imaging (MRI) studies have shown distinct cerebellar atrophy associated with LOTS. In this study, we furthered this investigation to determine if the cerebellar atrophy is globally uniform or preferentially targets certain cerebellar regions. We utilized *DeepCERES*, a deep learning cerebellar specific segmentation and cortical thickness pipeline to analyze differences between LOTS (n=20), LOSD (n=5), and neurotypical controls (n=1038). LOTS had smaller volumes of the whole cerebellum as well as cerebellar lobules IV, V, VI, VIIB, VIIIA, VIIIB, IX, and both Crus I and II compared to both LOSD and neurotypical controls. LOTS patients also had smaller cortical thickness of cerebellar lobules V, VI, VIIB, VIIIA, VIIIB, and both Crus I and II compared to both LOSD and neurotypical controls. Cerebellar functional and lesion localization studies have implicated lobules V and VI in speech articulation and execution while lobules VI, Crus I, VIIA, among others, have been implicated in a variety of behaviors and neuropsychiatric symptoms. Our observations provide a possible anatomical substrate to the higher prevalence of dysarthria and psychosis in our LOTS but not LOSD patients. Future studies are needed for direct comparisons considering phenotypic aspects such as age of symptom onset, presence and severity of dysarthria and ataxia, full characterization of neuropsychiatric profiles, molecular pathology and biochemical differences to fully understand the dichotomy observed in these two diseases.

## Introduction

The GM2 gangliosidoses are a group of ultra rare progressive autosomal recessive lysosomal storage disorders [1]. The GM2 gangliosidoses can be divided into three separate diseases based on the specific genes causing the pathological accumulation of the GM2 ganglioside in lysosomes leading to a cascade of events ultimately resulting in neurodegeneration [2,3]. Tay-Sachs disease, the most common form, is caused by biallelic variants in *HEXA* encoding the α subunit of the heterodimeric enzyme β-hexosaminidase A [4,5]. Sandhoff disease is caused by biallelic variants in *HEXB* encoding the β subunit of β-hexosaminidase A [4,6]. GM2 activator deficiency, the rarest form of GM2 gangliosidosis, is caused by biallelic variants in *GM2A* encoding the GM2 activator protein required as a cofactor to β-hexosaminidase A in order to degrade GM2 ganglioside [7,8]. Tay-Sachs disease occurs in 1 in 200,000 to 1 in 320,000 individuals with higher prevalence in some Brazilian and Ashkenazi Jewish populations due to founder effects [9–11]. Sandhoff disease occurs in 1 in 500,000 to 1 in 1,500,000 individuals.

GM2 activator deficiency has only 13 reported cases [8]. The diagnosis of the GM2 gangliosidoses has evolved over the last decades and involves the complementary use of β- hexosaminidase enzyme activity and molecular testing [12,13].

While GM2 gangliosidosis, like most lysosomal storage disorders is a systemic disease [14], the presence of large amounts of ganglioside in neuronal membranes with inability to fully degrade ganglioside, accounts for the high prevalence of central and peripheral nervous system dysfunction emerging from pathological GM2 substrate accumulation [15,16]. Symptom onset and disease progression in the GM2 gangliosidoses emerge along a continuum determined by the magnitude of residual β-hexosaminidase A enzyme activity [17,18]. For heuristic purposes, three clinical subtypes are recognized [18]. The infantile form has the least amount of residual enzyme activity and is the most severe with symptom onset occurring within the first 6 months of life [4]. These patients frequently present with halted development, abnormal startle response, hypotonia, and cherry red maculae [4]. Rapid decline occurs following symptom onset with death between 3 and 5 years of age [4,19]. The juvenile form of the disease has symptom onset between 2 and 10 years with a diagnosis at around 12 years of age [11,20]. Juvenile patients frequently present with dysarthria, difficulties ambulating, dystonia, and develop both incoordination and intellectual impairments [11,21,22]. Death occurs in juvenile GM2 patients frequently in the second decade, most commonly by respiratory infection [11]. The adult or late-onset form of GM2 gangliosides (LOGG) has the highest residual enzyme activity, has onset in the first or second decade, and has the most heterogeneity in clinical symptoms and rate of progression [18,23]. Initial symptoms often include lower limb weakness and signs of cerebellar cognitive affective syndrome (CCAS) including dysarthria, tremor, dystonia and neuropsychiatric symptoms [18,24,25]. Despite significant morbidity, LOGG might not alter life expectancy [12,26]. There are no approved therapies for the GM2 gangliosidoses and symptomatic management is all that is currently available [17,27]. However, investigations into gene therapy are underway in patients including the use of a mixed AAVrh8-HEXA and AAVrh8-HEXB dose, enabling the potential treatment of both Tay-Sachs and Sandhoff diseases [28].

Despite shared biochemical deficiency in β-hexosaminidase A, late-onset Tay-Sachs (LOTS) patients have been shown to have an increased prevalence of psychosis and mania symptoms, and late-onset Sandhoff disease (LOSD) patients appear to have an increased prevalence of sensory neuropathy [24]. Previous studies on the natural progression of LOGG have demonstrated a distinct neuroradiological cerebellar pathology in LOTS when compared to LOSD [29,30]. LOTS patients have shown cerebellar atrophy and associated 4^th^ ventricular enlargement when compared to LOSD through volumetric analysis [29,30]. Furthermore, one study investigating individual cerebellar lobule atrophy in LOTS patients found atrophy across the cerebellar lobules, with the most extensive atrophy in lobules V and VI followed by VII and VIII compared to controls [31].

Atrophy of the pons and superior cerebellar peduncle by MRI has also been reported in patients with advanced LOGG [32]. Magnetic resonance spectroscopy (MRS) has demonstrated that LOTS patients have shown higher concentrations of myo-inositol (mI) and lower concentrations of *N*-acetyl aspartate (NAA), creatinine (Cr), and glutamate-glutamine (Glx) in the cerebellum when compared to LOSD [30,33]. Furthermore, in our previous analysis utilizing diffusion tensor imaging (DTI) we found distinct reductions in fractional anisotropy (FA) and increases in mean diffusivity (MD), axial diffusivity (AD), and radial diffusivities (RD) in cerebellar white matter pathways of LOTS patients when compared to LOSD patients [5]. However, to the best of our knowledge, no study has performed a detailed analysis of cortical thickness at the cerebellar lobule level in LOGG.

The cerebellum has recently received increased attention particularly with recent investigations of its role in cognitive functions and behavior [34]. Anatomically, the cerebellum can be divided into three functional areas based on the input source including the cerebrocerebellum, the spinocerebellum, and the vestibulocerebellum [35,36]. Furthermore, the cerebellum can be divided into distinct lobules where lobules I-V make up the anterior portion of the cerebellum, lobules VI-IX and both Crus I and II make up the posterior portion of the cerebellum, and lobule X makes up the flocculonodular portion of the cerebellum [37,38]. We conducted a brief literature search (summarized in Table 1) focused on identifying the function of the individual cerebellar lobules from functional magnetic resonance imaging and lesion studies. Ataxia, as a motor symptom, was found to be localized to the anterior portion of the cerebellum extending onto lobule VI [38–40]. Dysarthria is localized to lobules V and VI [41,42]. Neuropsychiatric symptoms localized to the posterior cerebellum lobule VI [40], however studies in patients with psychosis have found Crus I to also be also implicated, demonstrating a significant role of the posterior cerebellum in behavior [43].

**Table 1.**
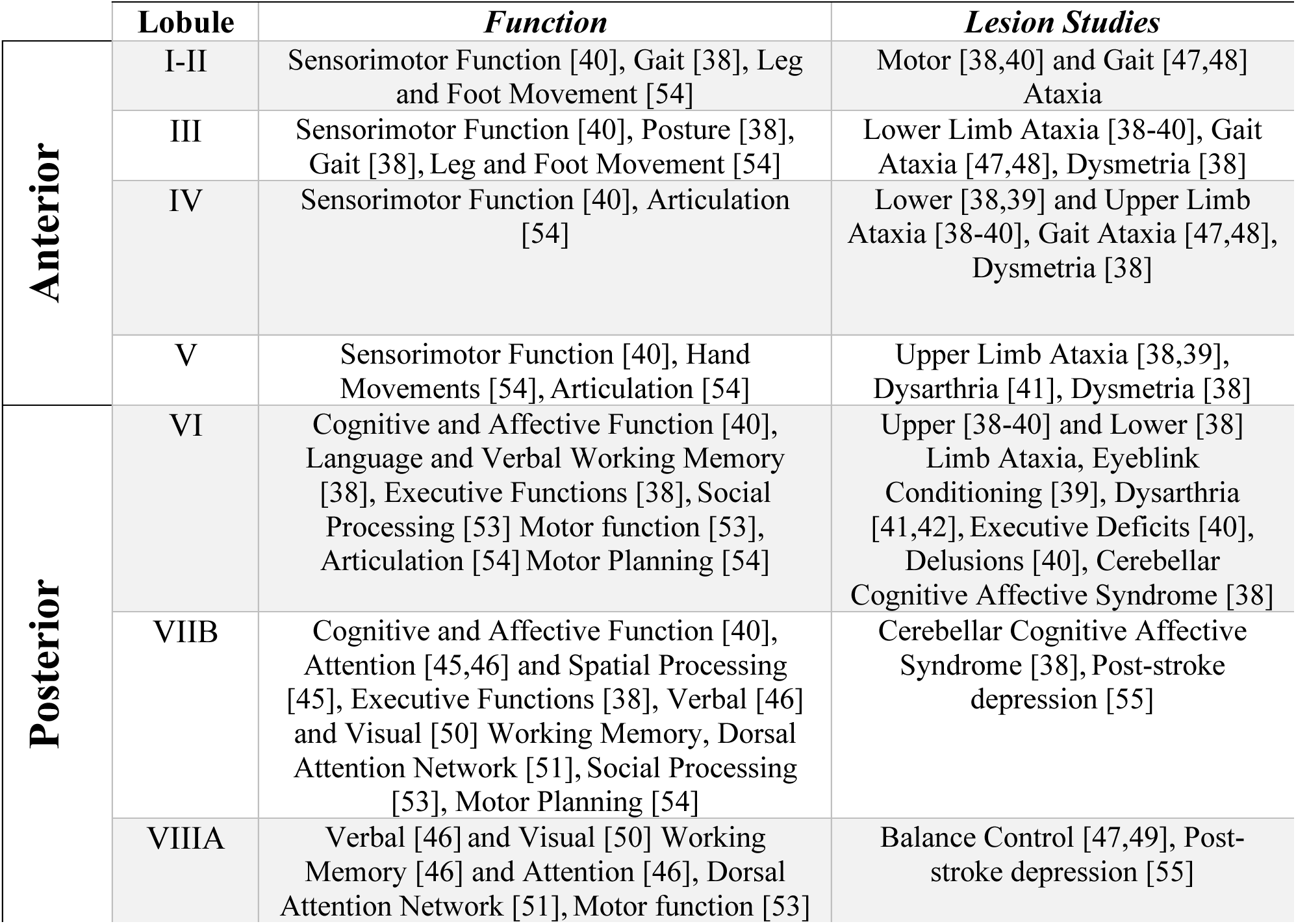
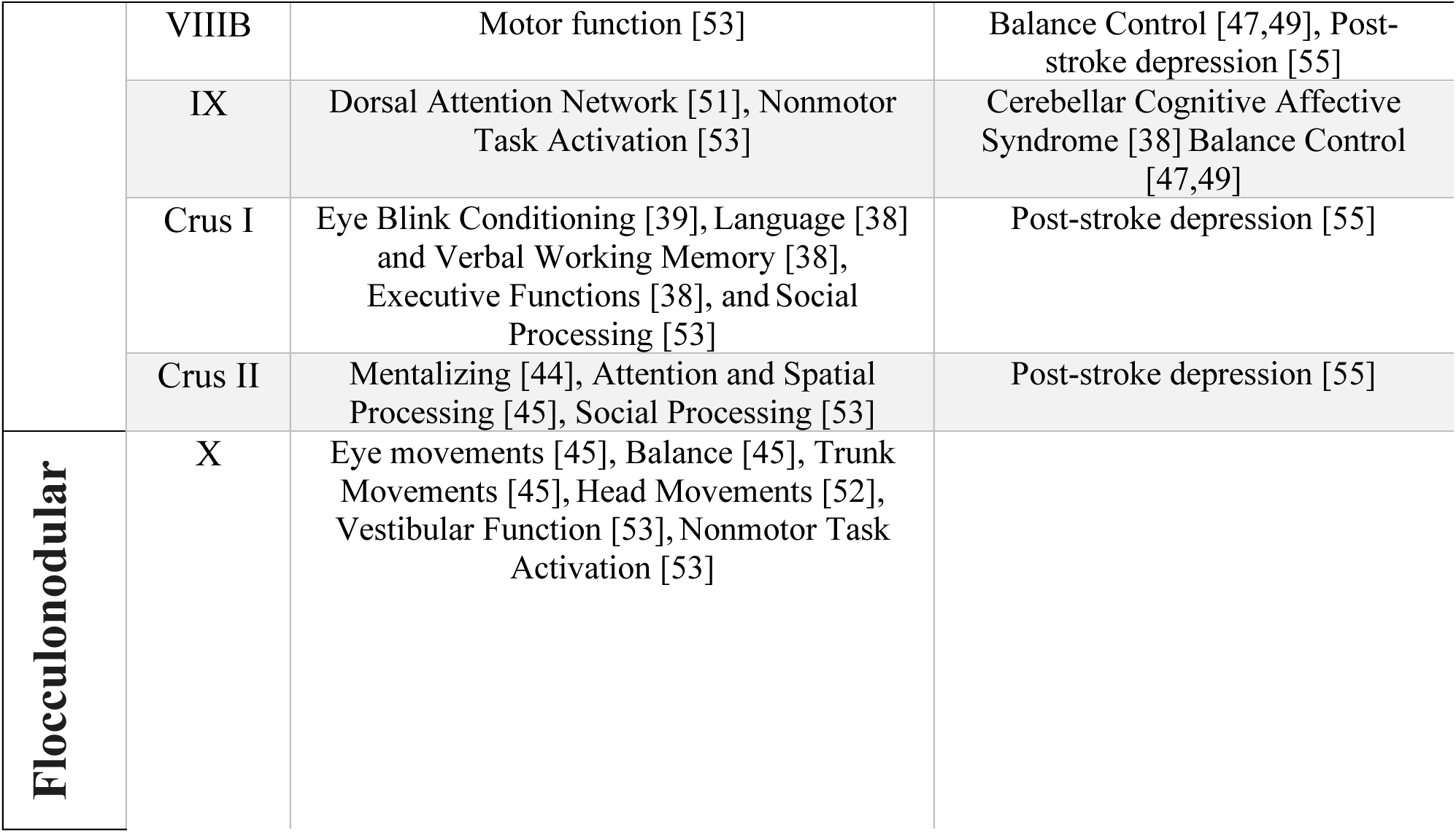
Cerebellar Lobule Function.

In this study, we sought to further define the magnetic resonance imaging natural history of LOGG by evaluating cerebellar lobule cortical thickness in patients with LOTS and LOSD compared to a pool of normative neurotypical controls. To the best of our knowledge, this is the first study evaluating cerebellar cortical thickness in GM2 gangliosidosis patients or a lysosomal storage disorder.

## Methods

### The Natural History of Late-onset GM2 Gangliosidosis (LOGG)

Participants from NCT00029965, the “Natural History of Glycosphingolipid & Glycoprotein Storage Disorders” with a diagnosis of either LOTS or LOSD and at least one T1-weighted MRI scan collected at the National Institutes of Health Clinical Center in Bethesda Maryland were included in this study. The NIH Institutional Review Board approved this protocol (02-HG- 0107). Informed consent was completed with all patients prior to participation and all research was completed in accordance with the Declaration of Helsinki. All patient visits were conducted at the National Institutes of Health Clinical Center (Bethesda MD) between 2010 and 2020. The distinction between LOTS and LOSD was made based on differences between β-hexosaminidase A and total β-hexosaminidase (β-hexosaminidase A + β-hexosaminidase B) activity patterns and biallelic variants in either *HEXA* or *HEXB* [5]. Twenty LOTS and five LOSD participants were included (Figure 1) and further information on LOTS and LOSD are provided in section A of the Supplementary Materials.

**Figure 1.**
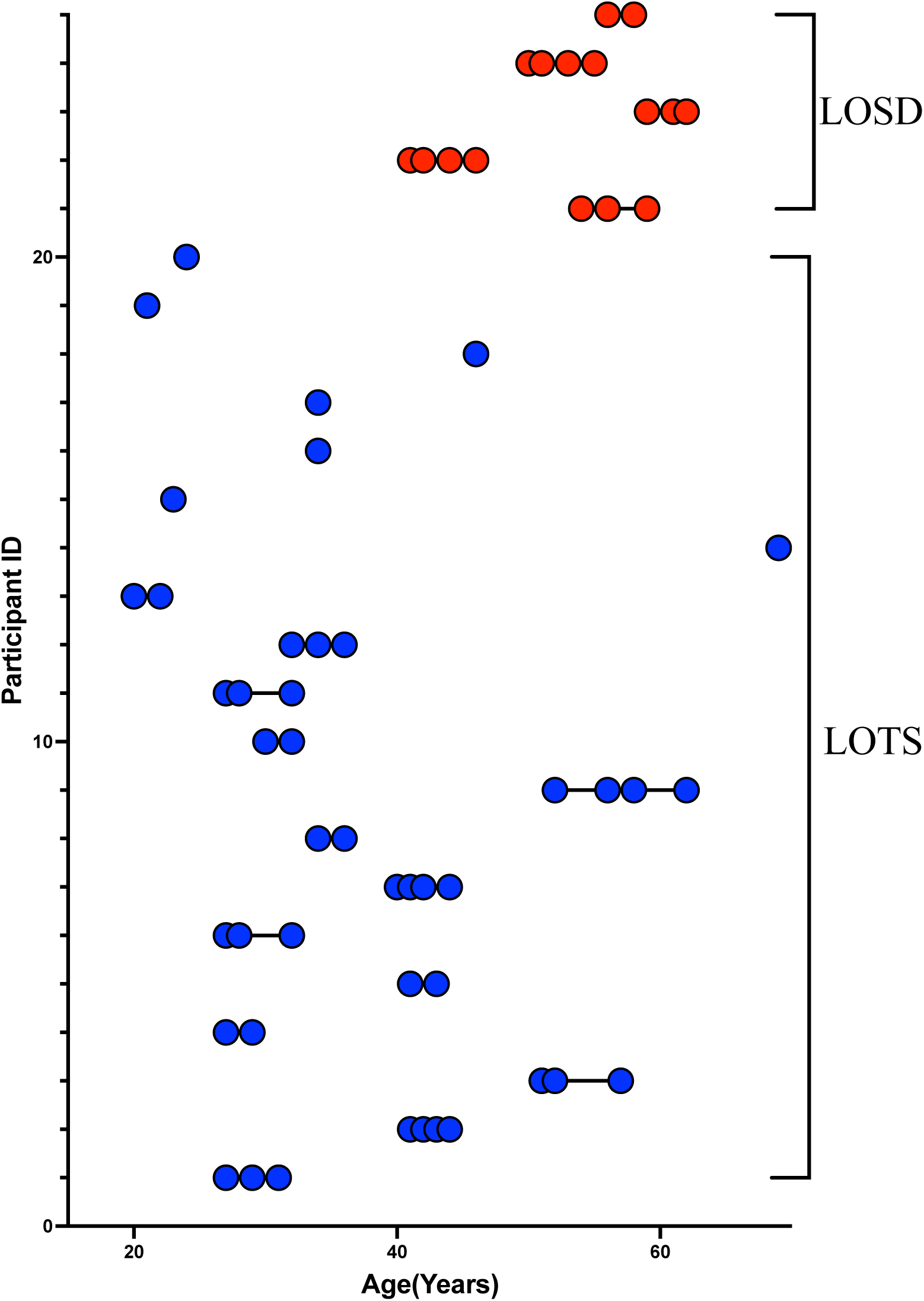
Participant age at each T1-weighted MRI Scan. Late-onset Tay-Sachs (LOTS, n = 20, 49 total MRI scans) patients are shown in blue and late-onset Sandhoff disease (LOSD, n = 5, 16 total MRI scans) patients are shown in red. Each T1-weighted scan is represented as a circle for all 65 scans where connecting lines indicate a repeated scan collected on the participant where each of the 25 participants are on a separate row.

### Neurotypical Controls

To account for the 51-year range in the LOGG cohort, we gathered 1038 neurotypical control MRI scans from four different open-source data repository datasets from OpenNeuro. One hundred eighty-seven participants from the “AgeRisk” dataset [56,57], 395 participants from the “Paingen_placebo” dataset [58,59], 155 participants from the “The NIMH intramural healthy volunteer dataset: A comprehensive MEG, MRI, and behavioral resource” [60,61], and 301 participants from the “Neurocognitive Aging Data Release with Behavioral, Structural, and Multi-echo functional MRI measures” [62,63] were included in this analysis. For more information on neurotypical control datasets, see section B of the Supplementary Materials.

### MRI Acquisition

Sixty-five MRI Scans from 25 LOGG patients were acquired on a Phillips Achieva 3T system with an 8-channel SENSE head coil. Sagittal MRI images were acquired with a 3D magnetization-prepared rapid acquisition with gradient echo (MPRAGE) sequence with the following parameters: TR/TE = 8/4 ms, slice thickness = 1 mm, flip angle = 8 degrees, NEX = 1, FOV = 220 mm.

### DeepCeres Volumetric and Cortical Thickness MRI Analysis

All sixty-five images from LOGG patients and 1038 neurotypical control scans were uploaded into volBrain’s deepCERES analysis pipeline. deepCERES is a deep learning analysis pipeline which was originally trained using 75 ultra high-resolution adult MRI images and later trained with 4,857 MRI images from various datasets reflecting a wide age range. deepCERES outputs volumes of the cerebellum including both gray and white matter, total intracranial volume (ICV), and bilateral volumes of cerebellar lobules I and II (combined), III, IV, V, VI, VIIB, VIIIA, VIIIB, IX, X, Crus I and Crus II which were all normalized to ICV (Figures 2 & 3). deepCERES also outputs average measured cortical thickness values of cerebellar lobules I and II (combined), III, IV, V, VI, VIIB, VIIIA, VIIIB, IX, X, Crus I and Crus II which were all normalized in relation to the cube root of ICV (adimensional).

**Figure 2.**
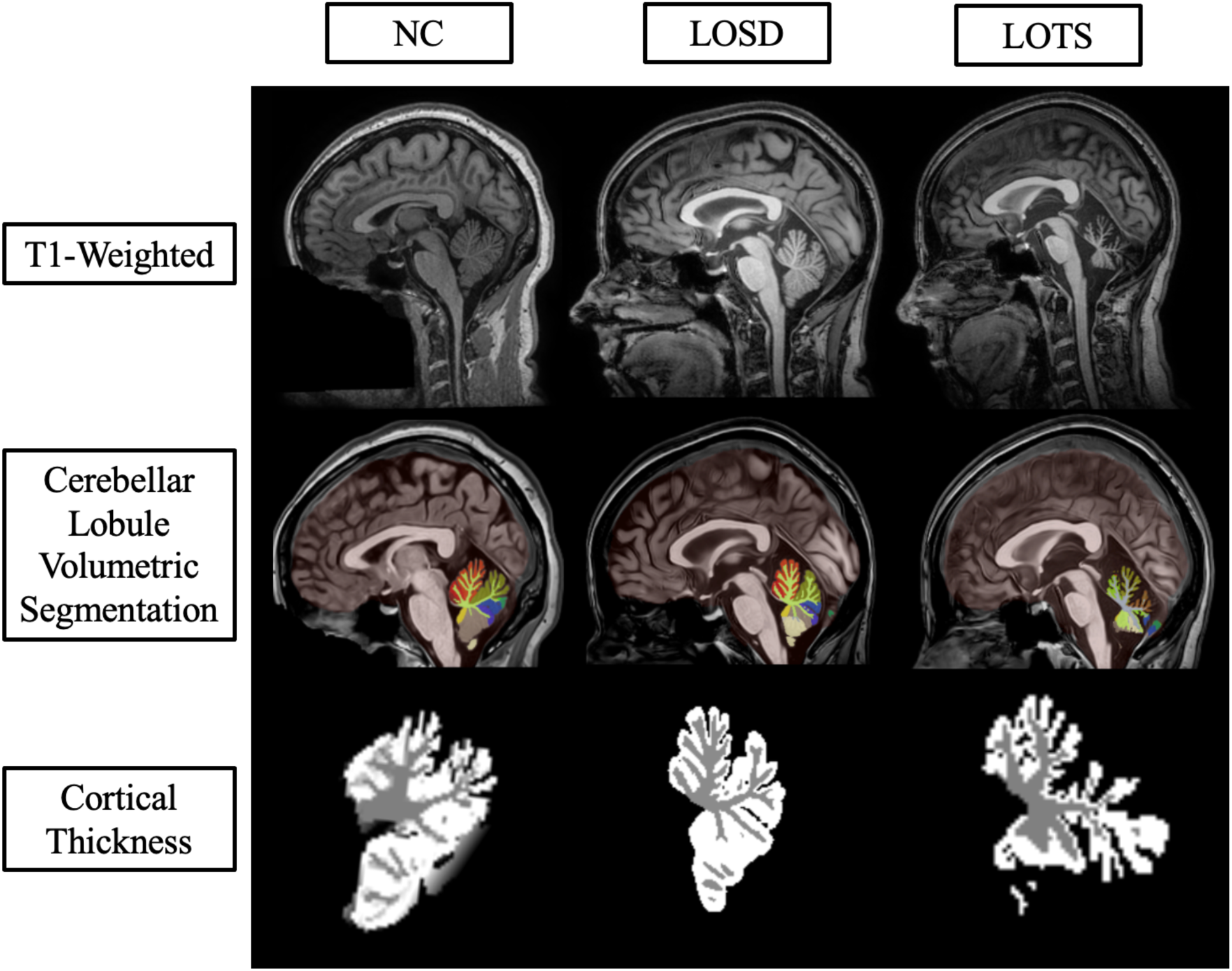
Sagittal deep learning segmentation of cerebellar lobules. Column one represents imaging from one 41- to 45-year-old neurotypical control (NC) female from the NIMH volunteer dataset. Column two represents imaging from one 41- to 45-year-old late-onset Sandhoff disease (LOSD) female patient. Column three represents imaging from one 41- to 45-year-old late-onset Tay-Sachs (LOTS) disease female patient. Row one represents unprocessed T1-weighted imaging for the three participants. Row two represents cerebellar lobule volumetric segmentation at the same slice for the three participants where distinct lobules are separated by color. Row three represents the cortical thickness estimation from *DeepCeres* at the same slice for the three participants where white matter is shown in grey and gray matter is shown in white. Axial and Coronal labels are shown in section C of the Supplementary Materials. Specific ages were redacted per Medrxiv requirements.

**Figure 3.**
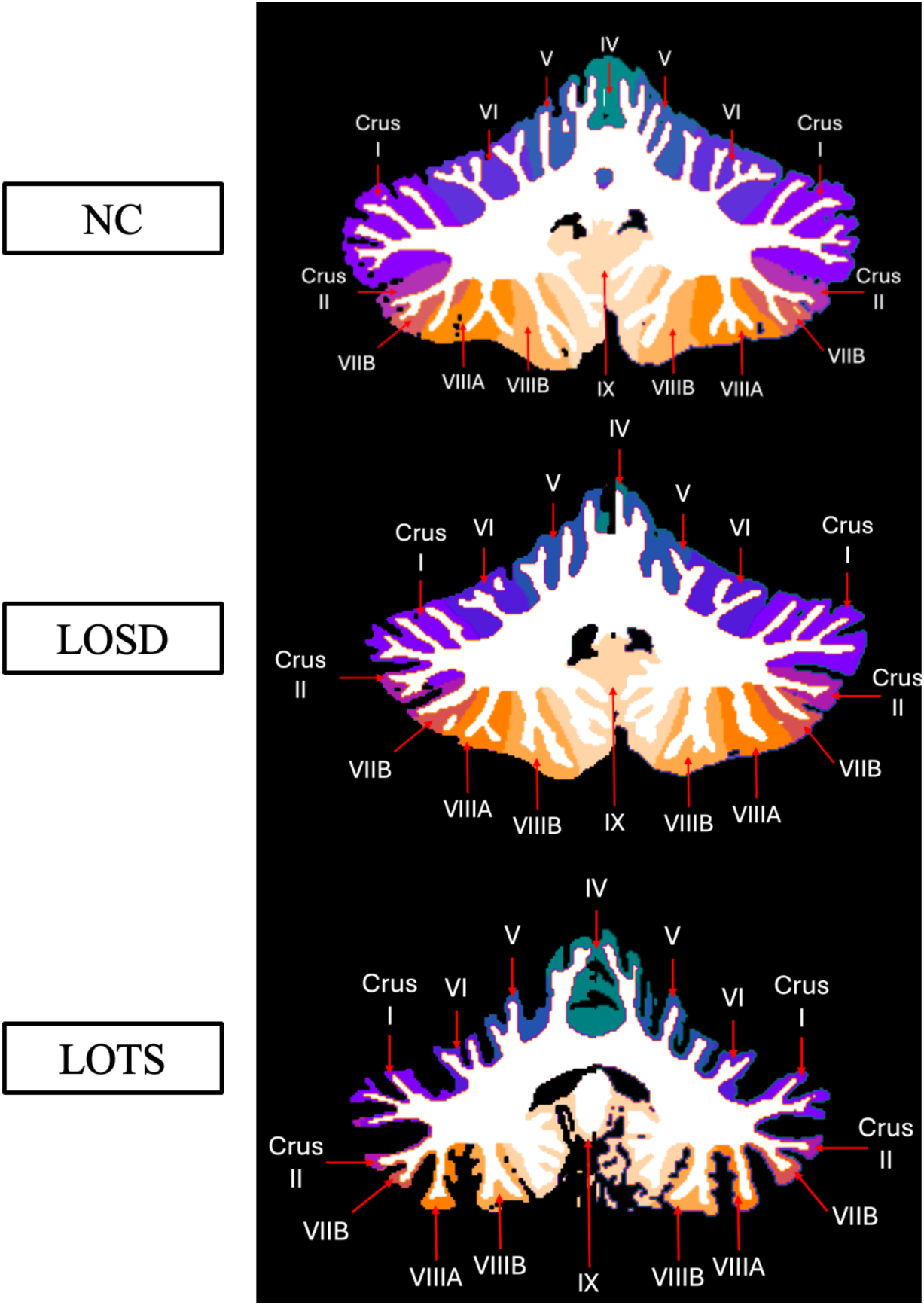
Labeled cerebellar lobule deep learning segmentation. Row one represents imaging from one 41- to 45-year-old neurotypical control (NC) female from the NIMH volunteer dataset. Row two represents imaging from one 41- to 45-year-old late-onset Sandhoff disease (LOSD) female patient. Row three represents imaging from one 41- to 45-year-old late-onset Tay-Sachs (LOTS) disease female patient. Each participants’ MRI scan was registered to the MNI space, and a coronal slice at coordinate y = -54 was shown for each participant. Sagittal and Coronal labels are shown in section D of the Supplementary Materials. Specific ages were redacted per Medrxiv requirements.

### Statistical Analysis

Statistical analysis in this study was performed in R (The R Foundation, v4.3.1) [64]. Between group analysis was performed using linear mixed effects models with age and sex (fixed effects) as a covariate of no interest to evaluate volumetric and cortical thickness data [5,65–67]. For comparisons between LOGG and NC, LOGG patients were assigned a value of 1 and NC were assigned a value of 0 to test the effects of LOGG. For comparisons between LOTS patients and NC, LOTS patients were assigned a value of 1 and NC were assigned a value of 0 to test the effects of LOTS disease. For comparisons between LOSD patients and NC, LOSD were assigned a value of 1 and NC were assigned a value of 0 to test the effects of LOSD. For comparisons between LOSD and LOTS, LOTS patients were assigned a value of 1 and LOSD patients were assigned a value of 0 to test distinctions in the two diseases. A subject level random intercept was used to account for repeated T1 scans conducted for each GM2 participant. *P-values* < 0.05 were designated as significant for volumetric and cortical thickness evaluations after a Bonferri correction for multiple comparisons. Volumetric values were controlled for ICV, and cortical thickness values were controlled in relation to the cube root of ICV.

## Results

### LOGG vs NC

LOGG patients, as a group, had smaller cerebellar volume (*χ*^2^(1) = 112.6, *p*-value_corrected_ < 0.0001) including both cerebellar gray matter (*χ*^2^(1) = 118.5, *p*-value_corrected_ < 0.0001) and cerebellar white matter (*χ*^2^(1) = 63.48, *p*-value_corrected_ < 0.0001) compared to NC when accounting for age and sex (Table 2). When analyzing specific cerebellar lobules, the volumes of lobules III, IV, V, VI, VI, VIIB, VIIIA, VIIIB, IX, and both Crus I and Crus II were smaller in LOGG patients compared to NC. Cerebellar lobule X volume was larger in LOGG patients compared to NC (*χ*^2^(1) = 7.52, *p*-value_corrected_ = 0.02). The combined volumes of cerebellar lobules I and II (*χ*^2^(1) = 0.43, *p*-value_corrected_ = 1.00) were not statistically different in LOGG patients compared to NC. The estimates and standard errors of the effect size between LOGG patients and NC are found in section G of the Supplementary Materials.

**Table 2.**
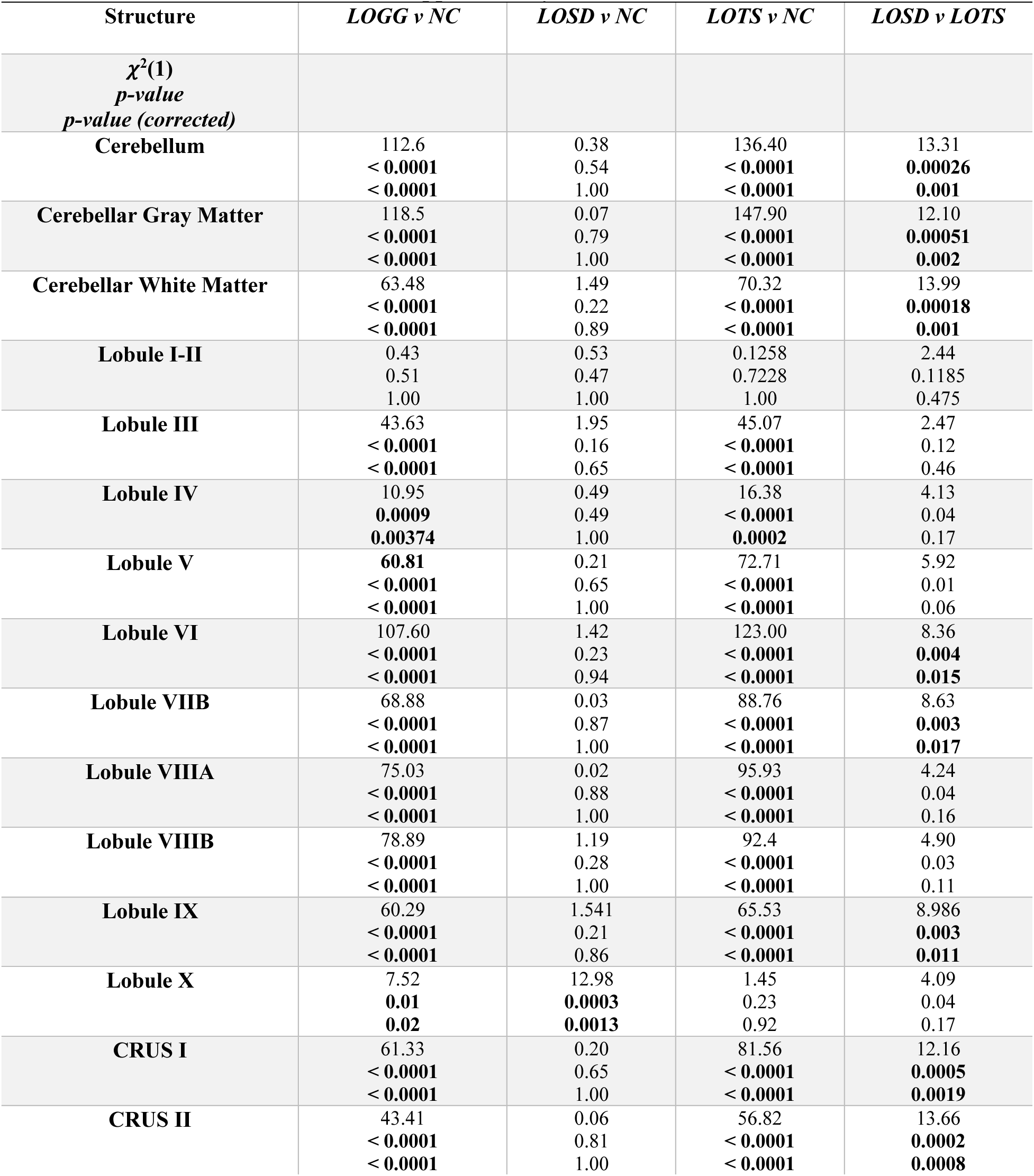
Cerebellar Volumetric MRI Analysis. All volumes were controlled for ICV. Statistical analysis was performed using a linear mixed effects model where *P*-values < 0.01 were considered significant and bolded. Estimates and Standard Error from the LMEM can be found in Supplementary Table G1.

Table 3 summarizes the results of the cortical thickness analysis. LOGG patients and NC. LOGG patients had smaller cerebellar cortical thickness in lobules I and II, III, V, VI, VIIB, VIIIA, VIIIB, IX, and both Curs I and Crus II compared to NC. There was no difference in the cerebellar lobule IV (*χ*^2^(1) = 0.01, *p*-value_corrected_ = 1.00) thickness between LOGG patients and NC. Lobule X cortical thickness was higher in LOGG patients compared to NC (*χ*^2^(1) = 13.86, *p*-value_corrected_ = 0.0008). The estimates and standard errors from cortical thickness analysis are shown in section H of the Supplementary Materials.

**Table 3.**
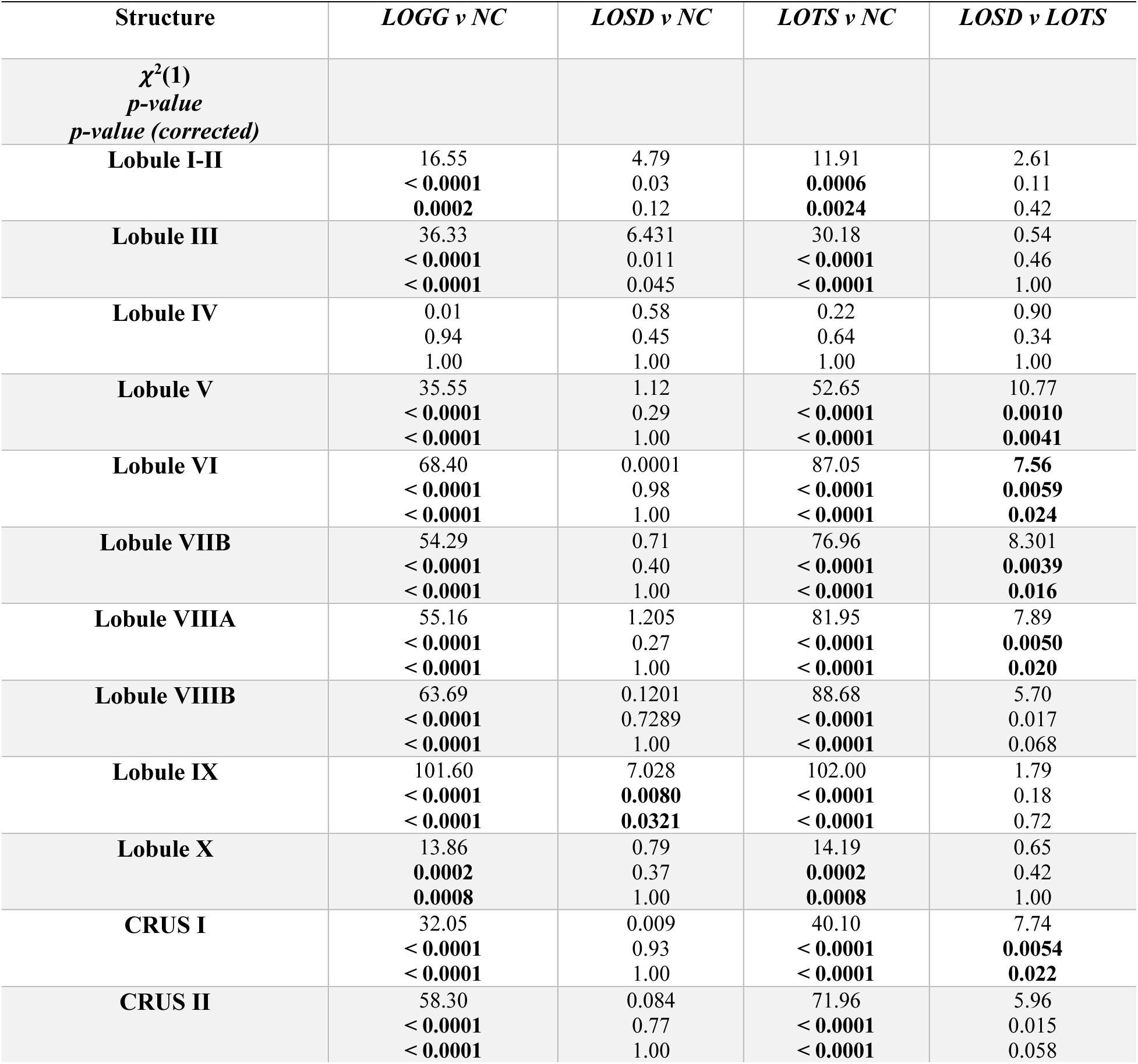
Cerebellar Cortical Thickness MRI Analysis. All volumes were all normalized in relation to the cube root of ICV (adimensional). Statistical analysis was performed using a linear mixed effects model where *P*-values < 0.01 were considered significant and bolded. Estimates and Standard Error from the LMEM can be found in Supplementary Table H1.

### LOTS vs NC

When LOTS patients were analyzed separately in comparison to NC, LOTS patients had significantly smaller cerebellar volume (*χ*^2^(1) = 136.40, *p*-value_corrected_ < 0.0001) including both gray matter (*χ*^2^(1) = 147.90, *p*-value_corrected_ < 0.0001) and white matter (*χ*^2^(1) = 70.32, *p*- value_corrected_ < 0.0001) compared to NC. LOTS patients also had significantly smaller cerebellar lobules III, IV, V, VI, VIIB, VIIIA, VIIIB, IX and both Crus I and II compared to NC (Table 2, Figure 4). There was no difference in cerebellar lobule volume between LOTS and NC in lobules I and II, and X.

**Figure 4.**
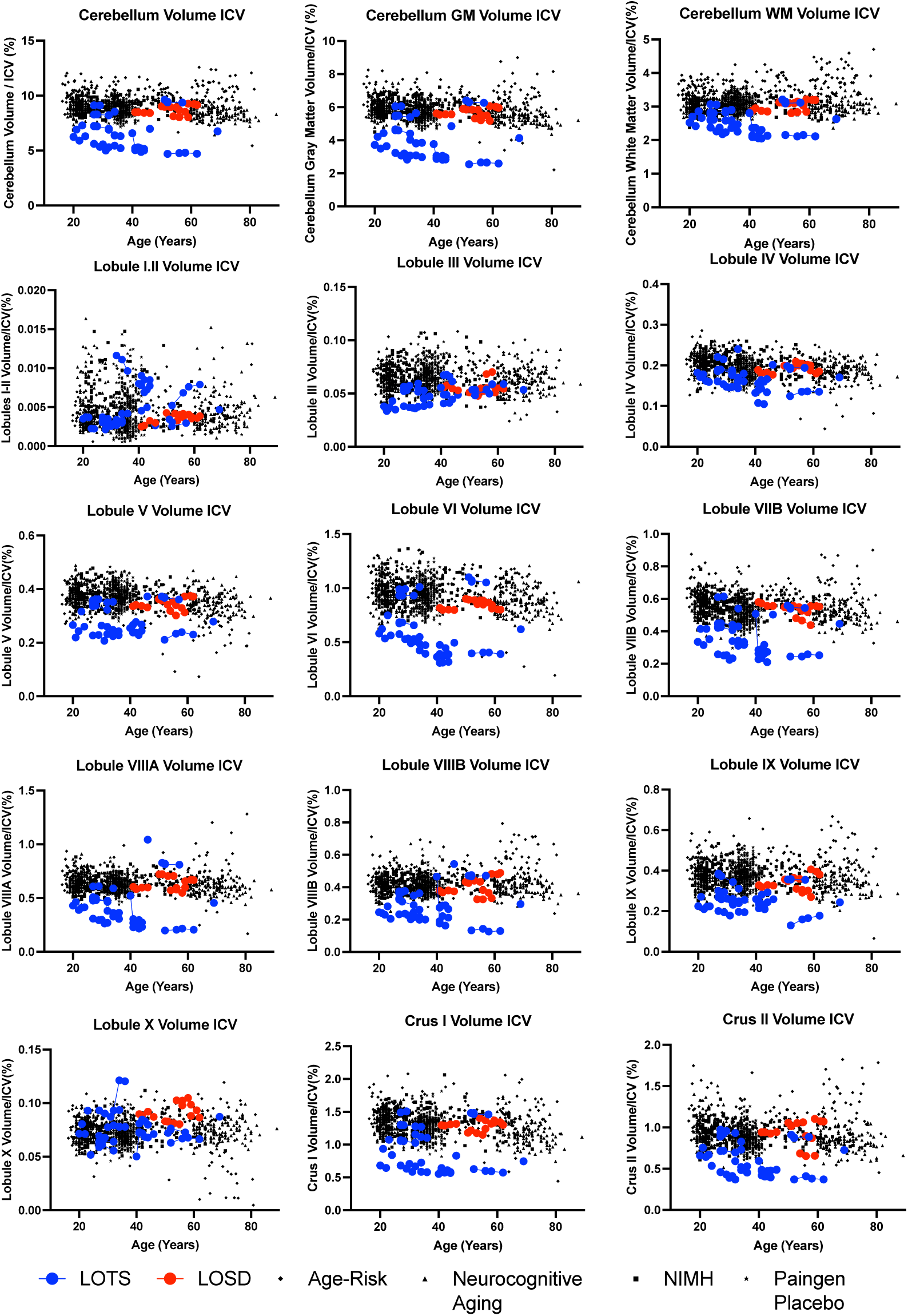
Volumetric Analysis of cerebellar lobule segmentation normalized to total intracranial volume (ICV). Neurotypical controls are shown as individual black points with different symbol shapes for each Open-Source data set. Late-onset Sandhoff Disease (LOSD) patients are shown as red circles with red connecting lines. Late-onset Tay-Sachs (LOTS) disease patients are shown as blue circles with blue connecting lines. Non-normalized values for volumetric analysis of the cerebellar lobules can be found in section E of the Supplementary Materials.

As summarized in Table 3, LOTS patients had smaller normalized cerebellar cortical thickness values for cerebellar lobules I and II, III, V, VI, VIIB, VIIIA, VIIIB, IX, and both Crus I and II when compared to NC. Lobule X (*χ*^2^(1) = 14.19, *p*-value_corrected_ = 0.0008) had higher normalized cerebellar cortical thickness in LOTS patients compared to NC (Table 3, Figure 5). There was no statistical difference between cerebellar lobule IV cortical thickness between LOTS patients and NC (*χ*^2^(1) = 0.22, *p*-value_corrected_ = 1.00).

**Figure 5.**
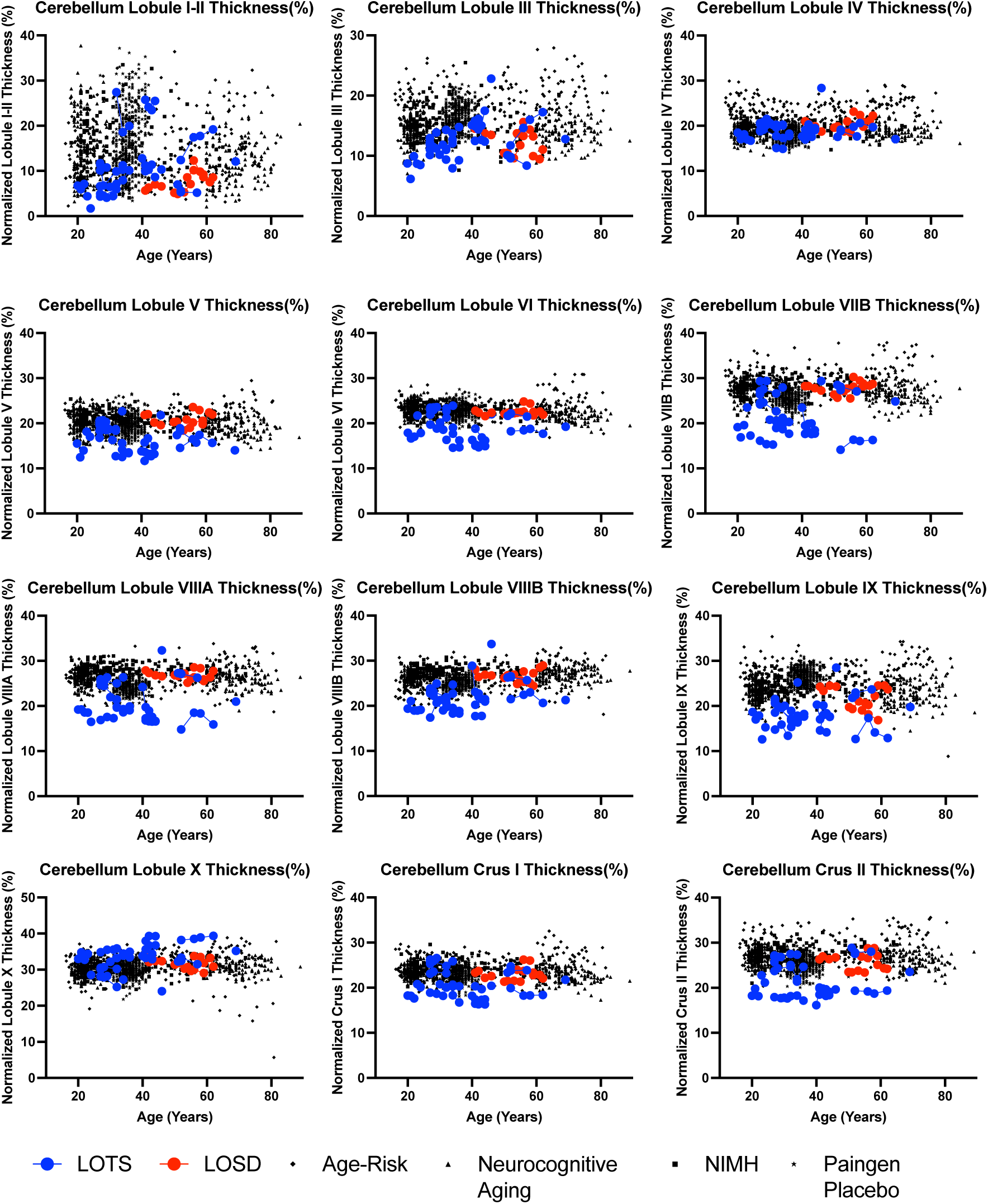
Cortical thickness analysis of the 12 cerebellar lobules normalized in relation to the cube root of ICV (adimensional). Neurotypical controls are shown as individual black points with different symbol shapes for each Open-Source data set. Late-onset Sandhoff Disease (LOSD) patients are shown as red circles with red connecting lines. Late-onset Tay-Sachs (LOTS) disease patients are shown as blue circles with blue connecting lines. Non-normalized values for cortical thickness of the 12 cerebellar lobules can be found in section F of the Supplementary Materials.

### LOSD vs NC

There was no difference in cerebellar volume including both cerebellar gray and white matter between LOSD patients and NC. When analyzing specific cerebellar lobules, LOSD patients had higher cerebellar lobule X volume (*χ*^2^(1) = 12.98, *p*-value_corrected_ = 0.0013) compared to NC. There was no difference in the volumes of cerebellar lobules I and II, III, IV, V, VI, VIIB, VIIIA, VIIIB, X, or either Crus I or II (*p*-value_corrected_ *>* 0.05, Table 2).

As shown in Table 3, LOSD patients had smaller cortical thickness in cerebellar lobules III (*χ*^2^(1) = 6.43, *p*-value_corrected_ = 0.045) and IX (*χ*^2^(1) = 7.03, *p*-value_corrected_ = 0.032) when compared to NC. No other cerebellar lobules had statistically different cerebellar cortical thickness differences between LOSD and NC (*p*-value_corrected_ *>* 0.05).

### LOTS vs LOSD

LOTS patients had statistically smaller cerebellum volume (*χ*^2^(1) = 13.31, *p*-value_corrected_ = 0.001) including both gray (*χ*^2^(1) = 13.99, *p*-value_corrected_ = 0.001) and white matter (*χ*^2^(1) = 12.10, *p*-value_corrected_ = 0.002) compared to LOSD patients. LOTS patients also had smaller volumes of cerebellar lobules IV, V, VI, VIIB, VIIIA, VIIIB, IX, X, and both Crus I and Crus II compared to LOSD patients. There was no statistical difference in the volumes of lobules I and II, and III between LOTS and LOSD patients (*p*-value_corrected_ *>* 0.05, Table 2).

As shown in Table 3, LOTS patients had smaller cortical thickness in lobules V, VI, VIIB, VIIIA, VIIIB, and Crus I. There were no other statistical differences observed between LOTS and LOSD patients for cortical thickness (*p*-value_corrected_ *>* 0.05).

## Discussion

Despite a long history of being indistinguishable and each respective gene encoding subunits of the same enzyme, recent studies have identified differences in the clinical presentation and trajectory of LOTS and LOSD. For instance, one study by Masingue et al. described an earlier symptom onset age and a higher prevalence of psychosis, dysarthria, and ataxia among LOTS patients with a higher prevalence of dysphagia in LOSD patients [24]. Numerous previous studies [30,68,69] have demonstrated cerebellar atrophy associated with LOGG, but until recently, none of these studies investigated the distribution of atrophy within the cerebellum. In our study, we used volBrain’s deepCERES, a deep learning cerebellar lobule segmentation algorithm to investigate both the volumes and cortical thickness of individual cerebellar lobules to provide further localization of this atrophy. In this study, we sought to further define the magnetic resonance imaging (MRI) natural history of LOGG.

LOGG patients were found to have significant cerebellar atrophy compared to controls including both cerebellar gray and white matter (Table 2). This was driven by LOTS patients as there was no statistical difference in cerebellar volume between LOSD and neurotypical controls.

Furthermore, the volumes of cerebellar lobules III, IV, V, VI, VIIB, VIIIA, VIIIB, IX, and both Crus I and II showed atrophy LOGG patients compared to controls (Table 2). This result was also exclusively driven by LOTS patients as there were no statistical differences observed between LOSD patients and controls for any of the lobules mentioned. When LOTS patients were compared directly to controls, the same lobules showed diminished volume. This result is consistent with the mouse model of LOTS which has showed GM2 storage across the cerebellum [70]. When LOTS patients were compared directly to LOSD patients, LOTS patients showed atrophy in the whole cerebellum (including both gray and white matter) and cerebellar lobules VI, VIIB, IX, and both Crus I and II demonstrating a distinct cerebellar pathology in LOTS compared to LOSD (Table 2, Figure 4).

The comparisons between LOTS patients and NC confirms many of the cerebellar results described in Májovská et al [31]. First, the most extensive cerebellar atrophy in LOTS patients compared to controls was found in lobules V, VI, VIIB, and Crus I with statistically significant atrophy observed in all cerebellar lobules analyzed in their study [31]. In our study, we found the most significant atrophy (*χ*^2^(1) > 50) in lobules V, VI, VIIB, VIIIA, VIIIB, IX, and both Crus I and II with statistically significant differences also observed in lobules III and IV. However, there were some distinctions. In the study by Májovská, lobules I-IV (the anterior cerebellum) was analyzed as a single structure [31]. Since the DeepCeres pipeline utilized in our study can segment out lobules III and IV individually from lobules I-II, and we observed cerebellar atrophy only in lobules III and IV in LOTS compared to NC, we suspect that atrophy they observed in combined lobules I-IV was driven by lobules III and IV as shown in this study [31].

Furthermore, they observed atrophy in Lobule X which we did not observe (*p*-value_corrected_ = 0.9152). One potential explanation for this is Lobule X’s resistance to neurodegeneration potentially resulting from heat shock protein HSP25 which has been previously described [71]. The dichotomy of the two studies requires further investigation. Furthermore, their study did not include any LOSD patients, where our study found significant divergence between the two diseases.

Evaluations of cerebellar cortical thickness showed LOGG patients had decreased cortical thickness in lobules I and II, III, V, VI, VIIB, VIIIA, VIIIB, IX, and both Crus I and II (Table 3, Figure 5) compared to controls. This result was similarly driven by LOTS patients, as apart from cerebellar lobule IX, no statistical differences were observed between LOSD patients and NC. There was significant overlap between cerebellar lobules showing cerebellar volume loss and decreased cortical thickness, however there was some divergence. When comparing LOGG and LOTS with NC, there were no statistical differences observed in volumetric comparisons in lobules I-II. Furthermore, lobule IV did show volumetric reductions in LOGG and LOTS compared to NC, however there were no statistically significant differences in the cortical thickness of lobule IV. Since there are strong correlations between volumetric and cortical thickness reductions of the cerebral cortex caused by aging [72], future investigations are needed to confirm this result and evaluate how this relates to clinical manifestations in these patients.

In the comparisons between LOTS and LOSD, reductions in cerebellar cortical thickness were observed in lobules V, VI, VIIB, VIIIA, VIIIB, and Crus I. This diverges from the volumetric analysis which found volumetric reductions in lobule IX and Crus II. However, volumetric reductions were not found in lobules V, VIIIA, and VIIIB where there were thickness reductions. This increased discrepancy could be caused by the reduced small sample size of the LOTS (n = 20) and LOSD (n = 5) cohorts, prompting further investigation. These cerebellar cortical thickness results provide a novel methodology for evaluating the cerebellar pathology in LOGG. Decreased cerebellar cortical thickness has previously been described in World Trade Center responders who had cognitive impairments, and was associated with episodic memory, response speed, and balance [73]. Furthermore, decreased cerebellar cortical thickness has also been associated with behavior symptoms in autistic spectrum disorder [74].

The importance of cerebellar dysfunction has received increased attention. As described in Table 1, cerebellar lesion studies have shown that dysfunction of lobules I-V of the anterior cerebellum are associated with ataxia. Dysfunction of lobules V and VI are associated with dysarthria, and dysfunction of lobule VI has been associated with delusions. In this study, we found overlap between these reports and cerebellar dysfunction in our LOGG patients. Furthermore, we provide further evidence towards the dichotomy between LOTS and LOSD as we found that LOTS patients had significantly smaller lobule V and VI volumes when compared to LOSD and NC. This supports the conclusion that cerebellar dysfunction in LOTS patients is the cause of the increased dysarthria present in these patients. Furthermore, cognitive and affective dysfunctions have been associated with alterations in cerebellar lobules VI and VIIB which may be a potential explanation for the observed increase of psychosis in LOTS patients.

Cerebellar Cognitive Affective Syndrome (CCAS) or Schmahmann Syndrome represents a profile of neurocognitive deficits including deficits in executive function, linguistic spatial cognition, and affect regulation emerging from cerebellar dysfunction [75,76]. Symptoms of CCAS have been documented in a small cohort of LOGG patients [25] and attributed to posterior cerebellar pathology including lobules VI, VII, and possibly lobule IX [76]. Based on the patterns of cerebellar atrophy in our larger patient cohort showing significant lobular and cortical cerebellar atrophy of lobules VI, Crus I, and VIIA, in LOTS patients compared to both NC and LOSD patients also supports the concept of CCAS being more prevalent in LOTS compared to LOSD patients.

There are limitations of this study which need to be considered before larger generalizations. First, this study is limited by a small sample size (n = 25), with five LOSD patients and twenty LOTS patients. While the rarity of LOTS and most notably LOSD limit large scale data collection, future studies could implement a quasi-study design like Masingue et al [24] which employed data from the literature to boost their sample size. From an imaging perspective, variations in MRI sites, scanners, and protocols are a limitation of this study. While all LOTS and LOSD patients were scanned on the same protocol, brand and type of scanner (3T Phillips Achieva), this was not the case for NC participants as they were scanned on Siemens or General Electric (GE) 3T scanners (more information on NC MRI acquisition is provided in Section B of the Supplementary Materials). However, we think this variance was mitigated since most of the NC participants were scanned at a high resolution (1 mm isotropic or better). Ultimately, future studies should strive for uniform scanning protocols across all participants. Lastly, the present study utilizes only T1-weighted imaging. Future studies are needed to make direct correlations between global and lobular cerebellar dysfunction, including multiple domains of MRI including DTI and MRS, with the potential involvement of functional magnetic resonance imaging (fMRI) to cement and fully define the link between cerebellar dysfunction and clinical implications in both LOTS and LOSD. For instance, functional networks involved in speech production [77], delusions [78,79], and motor function [80] should be evaluated in their relation to manifesting symptoms.

## Conclusion

In this study, we aimed to discern differences in cerebellar lobule volume and cortical thickness between LOGG patients, LOTS patients, LOSD patients, and neurotypical controls. To our knowledge, this is the first study using individual cerebellar lobule segmentation in LOGG to differentiate disease subtypes. Volumetric atrophy and cortical thickness reductions were observed across numerous cerebellar lobules in LOGG and LOTS patients compared to controls. LOSD patients only demonstrated reductions in cortical thickness in cerebellar lobule IX compared to controls. LOTS patients also demonstrated numerous lobules with volumetric atrophy and cortical thickness reductions compared to LOSD patients which is consistent with previous studies describing a higher prevalence of cerebellar deficits in LOTS.

Furthermore, through our literature search, we found disruption of lobules V and VI to be associated with dysarthria and lobules VI, Crus I, VIIA, among others, to be associated with a variety of cognitive function and neuropsychiatric deficits. Combined with the localization of atrophy in LOTS patients, this study provides further evidence towards the notion that dysfunction in these lobules could represent, at least in part, an anatomical substrate for dysarthria and psychosis in LOTS. However, future studies should investigate the cerebellar metabolic dichotomy between LOSD and LOTS, and to correlate clinical symptoms with cerebellar atrophy in LOTS to anchor these findings.

## Acknowledgements

We thank the participants and their families for the generosity of their time and efforts. We are also grateful to many staff members and care providers who contributed their expertise over the years.

## Data Availability

The data described in this manuscript are available from the corresponding author upon reasonable request. Neurotypical control data are available from OpenNeuro at the following links; AgeRisk: https://openneuro.org/datasets/ds004711/versions/1.0.0 [56,57], Paingen_Placebo: https://openneuro.org/datasets/ds004746/versions/1.0.1 [58,59], NIMH: https://doi.org/10.18112/openneuro.ds004215.v1.0.0 [60,61], Neurocognitive Aging Data Release with Behavioral, Structural, and Multi-echo functional MRI measures: https://openneuro.org/datasets/ds003592/versions/1.0.13 [62,63].

## Author Contributions

Conceptualization: CJL, SIC, MTA, CT, CJT; Data Curation: JMJ, CJT, MTA; Funding Acquisition: CJT; Methodology: CJL, SIC, JMJ, CT, MTA, CJT; Visualization: CJL, SIC, CT, MTA, CJT; Writing-original draft: CJL, CJT; Writing-reviewing & editing; CJL, SIC, JMJ, CT, MTA, CJT

## Ethics Declaration

The NIH Institutional Review Board approved this protocol (02-HG-0107). Informed consent was completed with patients before participation and all research was completed in accordance with the Declaration of Helsinki.

## Abbreviations

AD: Axial Diffusivity
CCAS: Cerebellar Cognitive Affective Syndrome
Cr: Creatine
DTI: Diffusion Tensor Imaging
DWI: Diffusion Weighted Imaging
FA: Fractional Anisotropy
fMRI: Functional Magnetic Resonance Imaging
Glx: Glutamate-Glutamine
ICV: Intracranial Volume
LOGG: Late-onset GM2 gangliosidosis
LOSD: Late-onset Sandhoff Disease
LOTS: Late-onset Tay-Sachs
MCP: Middle Cerebellar Peduncle
MD: Mean Diffusivity
mI: myoinositol
MRI: Magnetic Resonance Imaging
MRS: Magnetic Resonance Spectroscopy
NAA: N-acetyl aspartate
NC: Neurotypical Control
QA: Quantitative Anisotropy
RD: Radial Diffusivity
SCP: Superior Cerebellar Peduncle

## Supplementary Methods

### Supplement A: Natural History Study Participant Characteristics

**Table A1.**
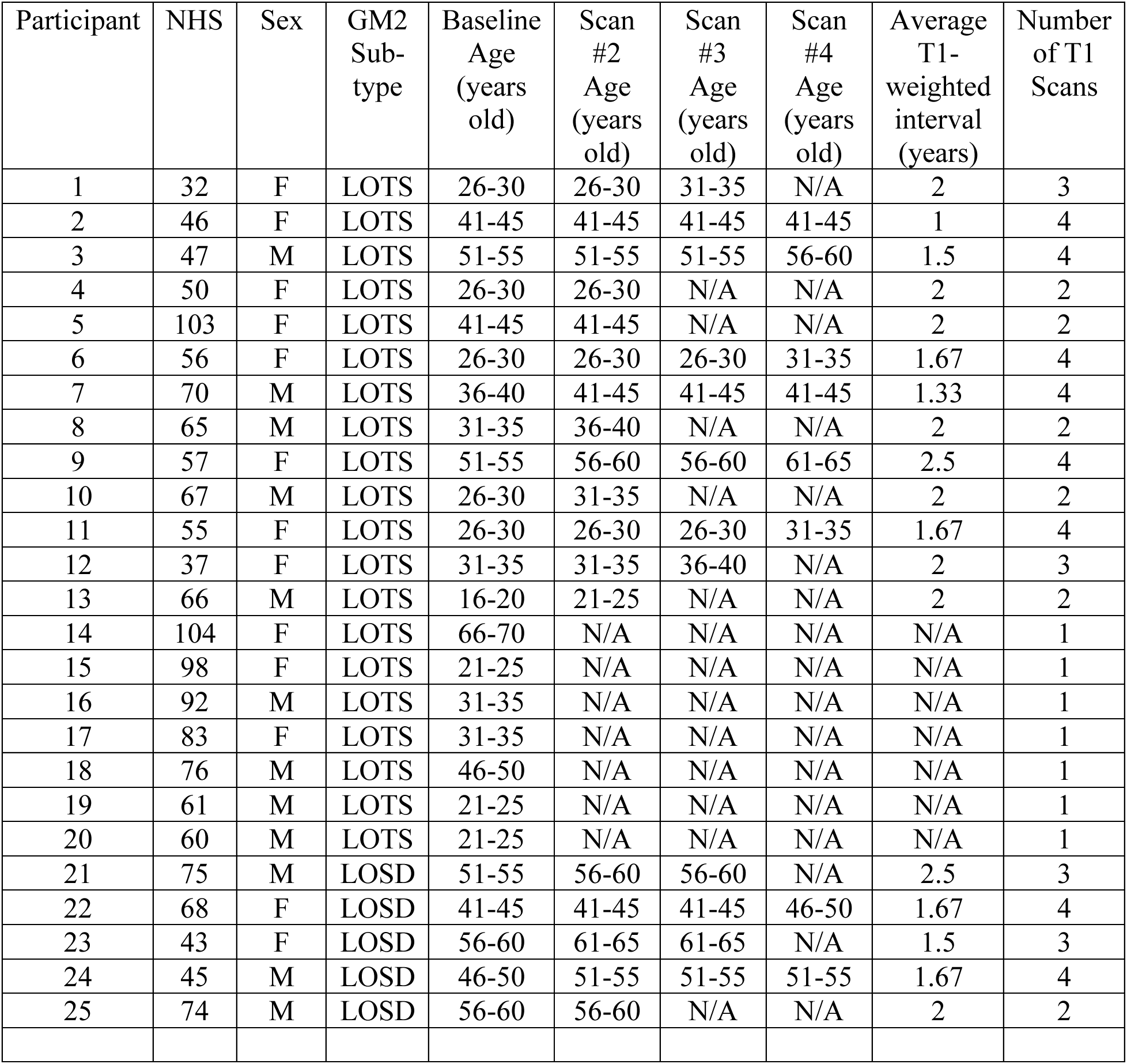
Natural History Study T1-Weighted MRI Scan Age (n = 25). N/A are designated when a participant did not have the above referenced MRI scan. Specific ages were redacted per Medrxiv requirements.

### Supplement B: Neurotypical Control Participant Characteristics

#### NIMH^2,3^

Participants from “The National Institute of Mental Health (NIMH) Intramural Healthy Volunteer Dataset”^2,3^ were scanned using a General Electric (GE) discovery MR750W 3T system with a 32-channel head coil. There were 62 participants with the FSPGR sequence and 92 participants with the MPRAGE sequence.^2,3^ With the following parameters:

**MPRAGE**
TR/TE = 6.95/2.92 ms
Flip angle = 8^∘^
Voxel = 1 mm isotropic voxels

**FSPGR**
TR/TE = 7.35/3.04 ms
Flip angle = 11^∘^
Slice Thickness = 1.2 mm

#### Neurocognitive Aging^4,5^

Participants from the “Neurocognitive aging data release with behavioral, structural, and multi-echo functional MRI measures” were included in this analysis.^4,5^ MRI data was acquired from one of two sites either the Cornell Magnetic Resonance Imaging Facility in Ithaca, New York or the York University Neuroimaging Center in Toronto, Canada^5^. MRI data from the Cornell University site was acquired on a 3T GE discovery MR750W 3T system with a 32-channel head coil. T1 weighted imaging was acquired with the following parameters: TR/TE = 2530/3.4 ms; 7° flip angle; 1 mm isotropic voxels, and 176 slices. MRI data from York University site was acquired on a 3T Siemens TimTrio MRI scanner with a 32-channel head coil. T1 weighted imaging was acquired from this site with the following parameters: TR/TE = 1900/2.52 ms, flip angle = 9°, 1 mm isotropic voxels, 192 slices.

#### Paingen_placebo^6,7^

Participants from the “Paingen_Placebo” data set were included in this analysis. MRI data was acquired on a Siemans Prisma 3T system with a 32-channel head coil at the University of Colorado at Boulder.^6^ T1 weighted imaging was acquired with the following parameters TR/TE = 2000/2.11 ms, flip angle = 8°, FOV = 256 mm, and resolution = 0.8 × 0.8 × 0.8 mm.

#### AgeRisk^8,9^

MRI data from the “AgeRisk” data set was acquired using a Siemens 3T MAGNETOM Prisma magnetic resonance imaging (MRI) system and a 20-channel head coil at the University Hospital Basel, Switzerland. T1-weighted MRI data was acquired using a magnetization-prepared rapid gradient echo sequence with the following parameters: TR/TE= 2500/4.25 ms, inversion time = 1100 ms, flip angle = 7^∘^, field of view = 256 mm × 256 mm, 192 slices, voxel dimensions = 1.0 mm isotropic.

**Figure B1.**
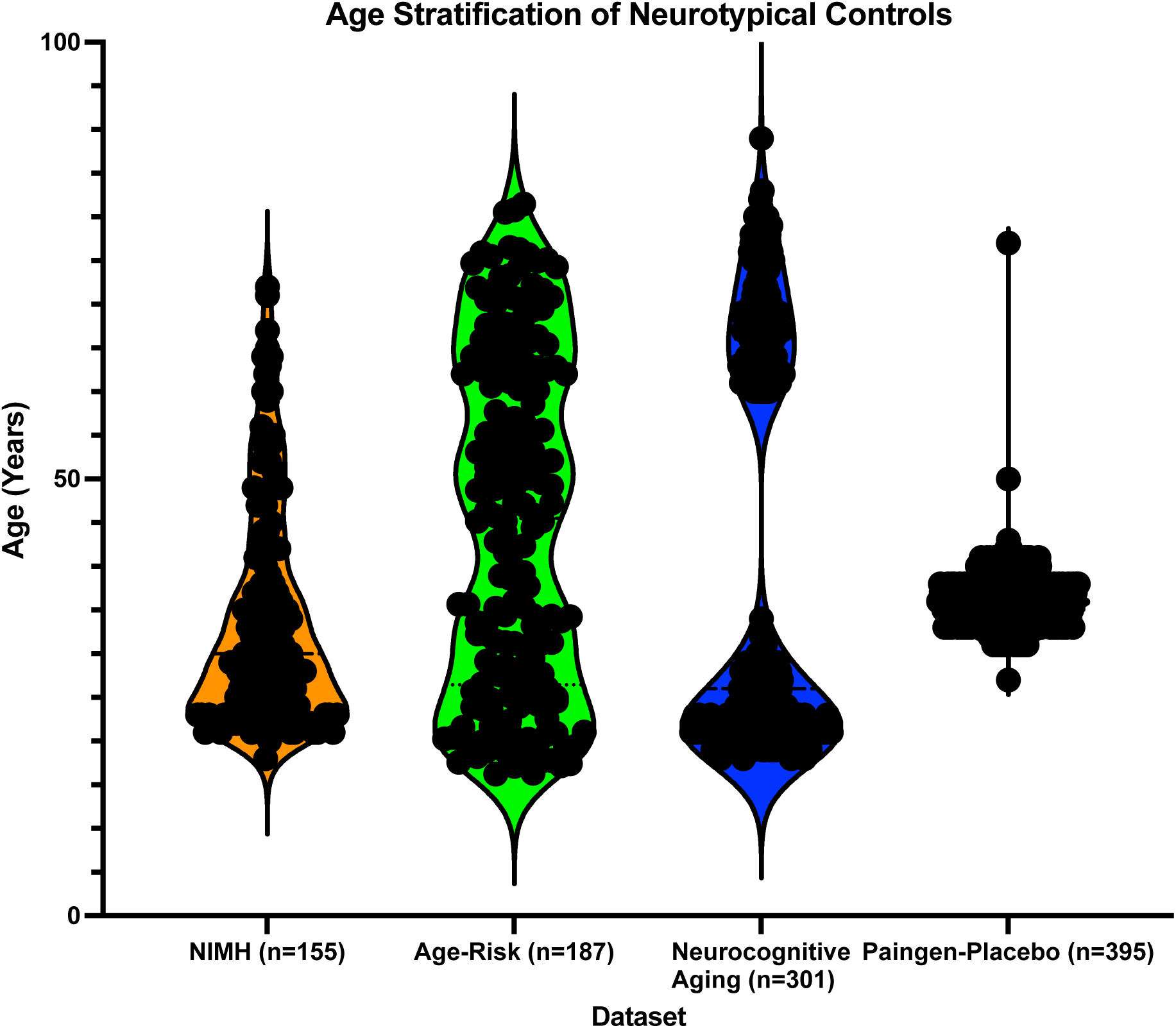
Violin Plot of Neurotypical Controls Age.

### Supplement C: Cerebellar Lobule Segmentation and Cortical Thickness

**Figure C1.**
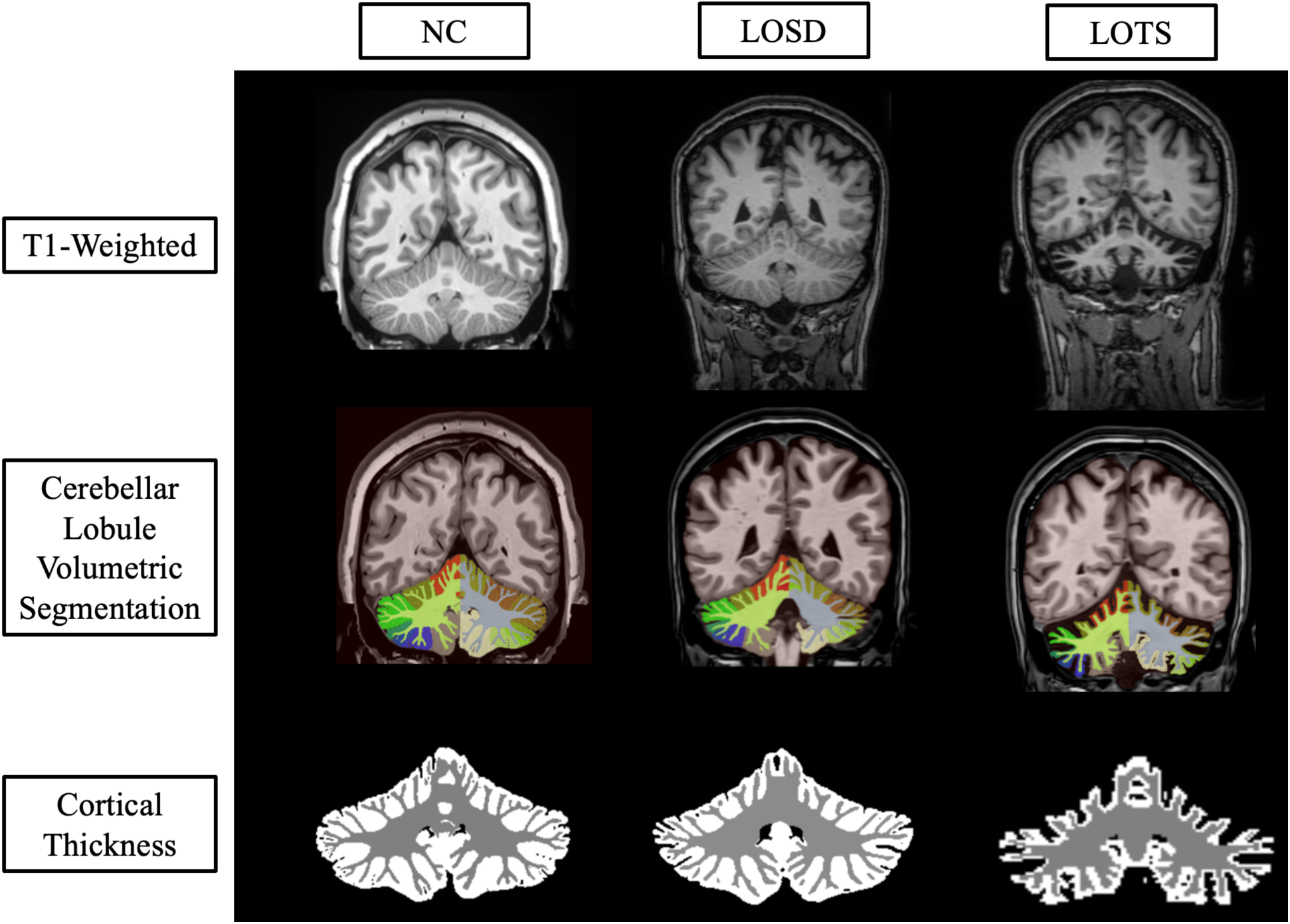
Coronal deep learning segmentation of cerebellar lobules. Column one represents imaging from one 41- to 45-year-old neurotypical control (NC) female from the NIMH volunteer dataset. Column two represents imaging from one 41- to 45-year-old late-onset Sandhoff disease (LOSD) female patient. Column three represents imaging from one 41- to 45-year-old -year-old late-onset Tay-Sachs (LOTS) disease female patient. Row one represents unprocessed T1- weighted imaging for the three participants. Row two represents cerebellar lobule volumetric segmentation at the same slice for the three participants where distinct lobules are separated by color. Row three represents the cortical thickness estimation from *DeepCeres* at the same slice for the three participants where white matter is shown in grey and gray matter is shown in white. Specific ages were redacted per Medrxiv requirements.

**Figure C2.**
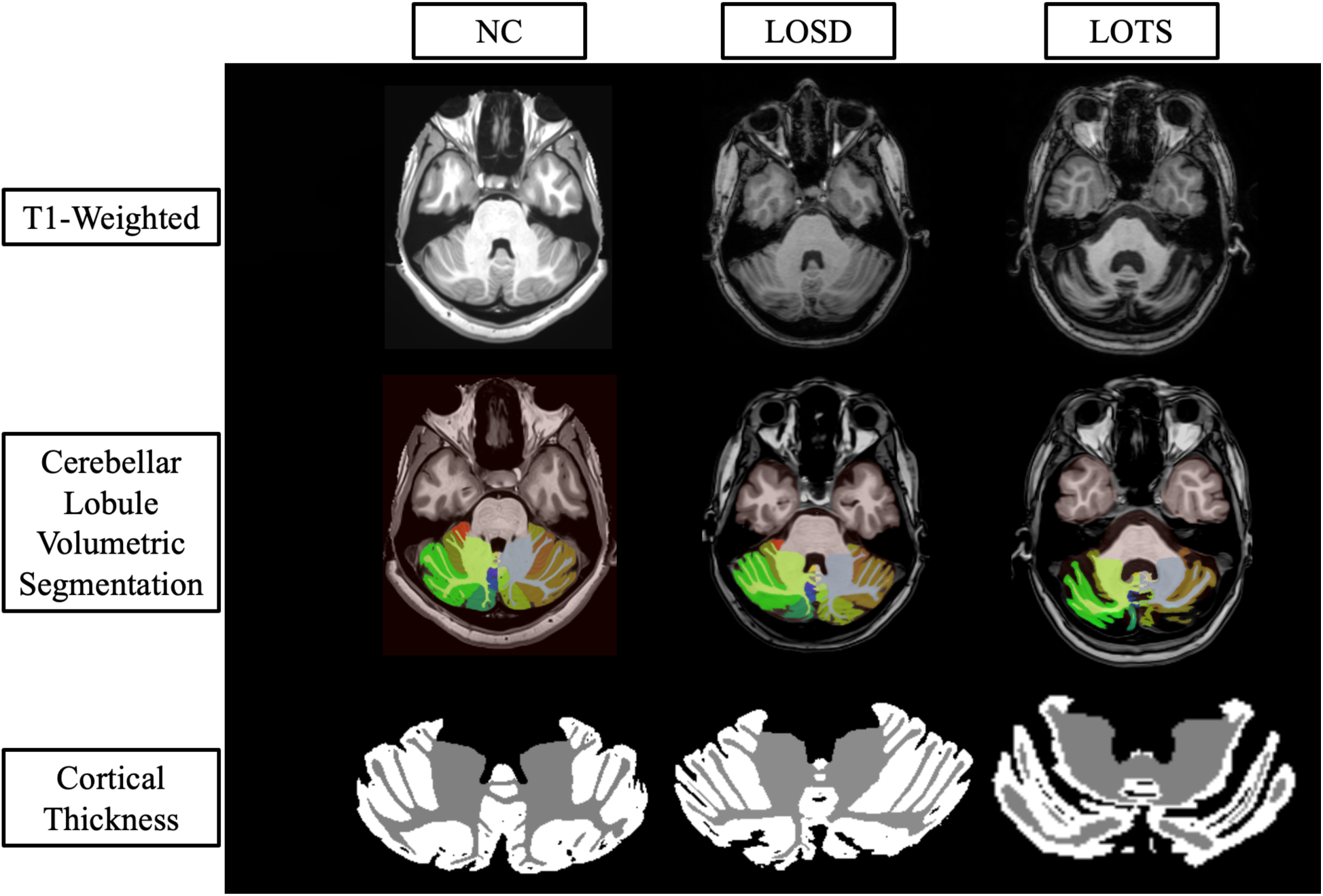
Axial deep learning segmentation of cerebellar lobules. Column one represents imaging from one 41- to 45-year-old neurotypical control (NC) female from the NIMH volunteer dataset. Column two represents imaging from one 41- to 45-year-old late-onset Sandhoff disease (LOSD) female patient. Column three represents imaging from one 41- to 45-year-old late-onset Tay-Sachs (LOTS) disease female patient. Row one represents unprocessed T1-weighted imaging for the three participants. Row two represents cerebellar lobule volumetric segmentation at the same slice for the three participants where distinct lobules are separated by color. Row three represents the cortical thickness estimation from *DeepCeres* at the same slice for the three participants where white matter is shown in grey and gray matter is shown in white. Specific ages were redacted per Medrxiv requirements.

### Supplement D: Cerebellar Lobule Individual Segmentation

**Figure D1.**
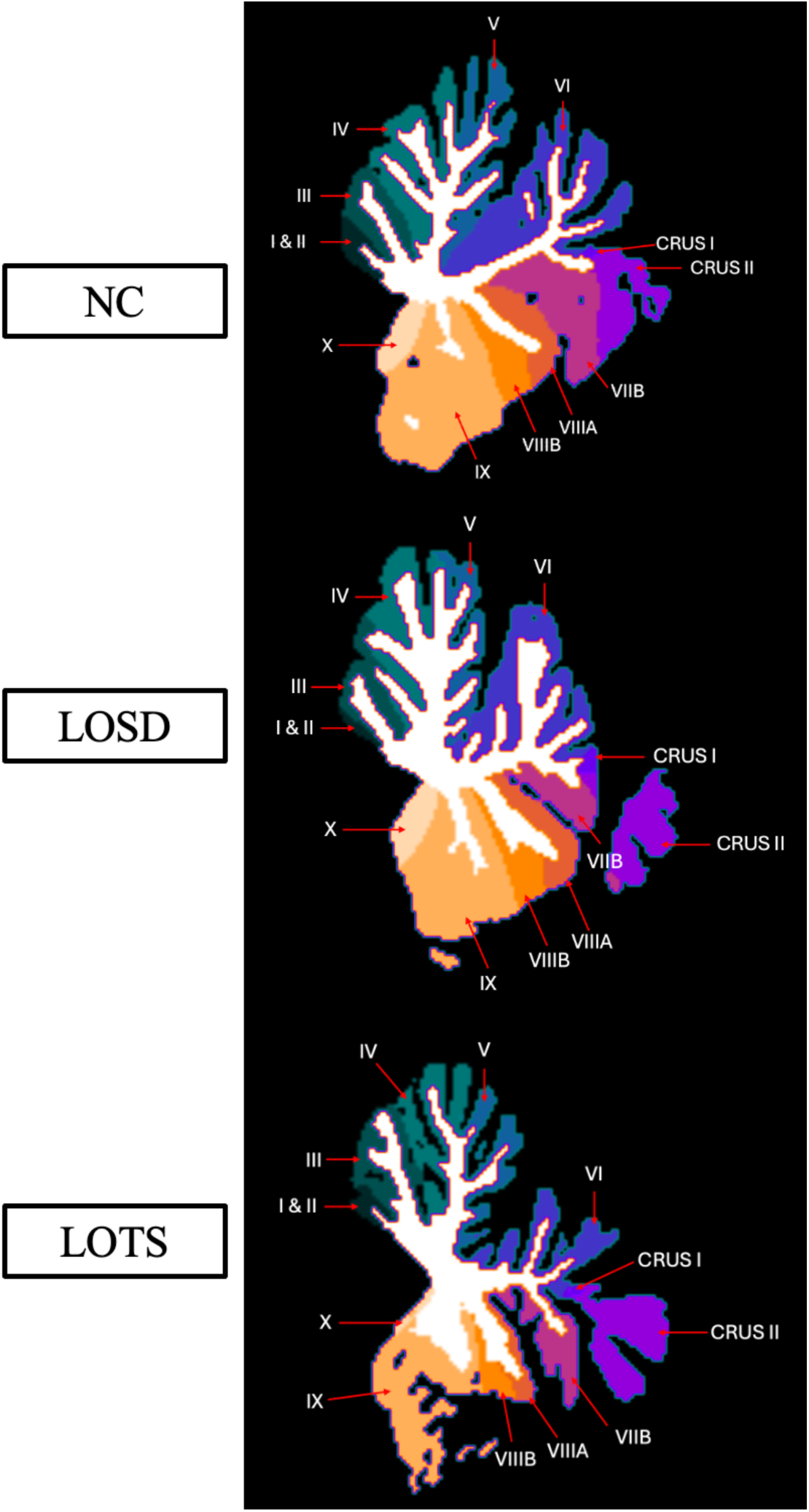
Labeled cerebellar lobule deep learning segmentation. Row one represents imaging from one 41- to 45-year-old neurotypical control (NC) female from the NIMH volunteer dataset. Row two represents imaging from one 41- to 45-year-old late-onset Sandhoff disease (LOSD) female patient. Row three represents imaging from one 41- to 45-year-old late-onset Tay-Sachs (LOTS) disease female patient. Each participants’ MRI scan was registered to the MNI space, and a sagittal slice at coordinate x = -2.5 was shown for each participant. Specific ages were redacted per Medrxiv requirements.

**Figure D2.**
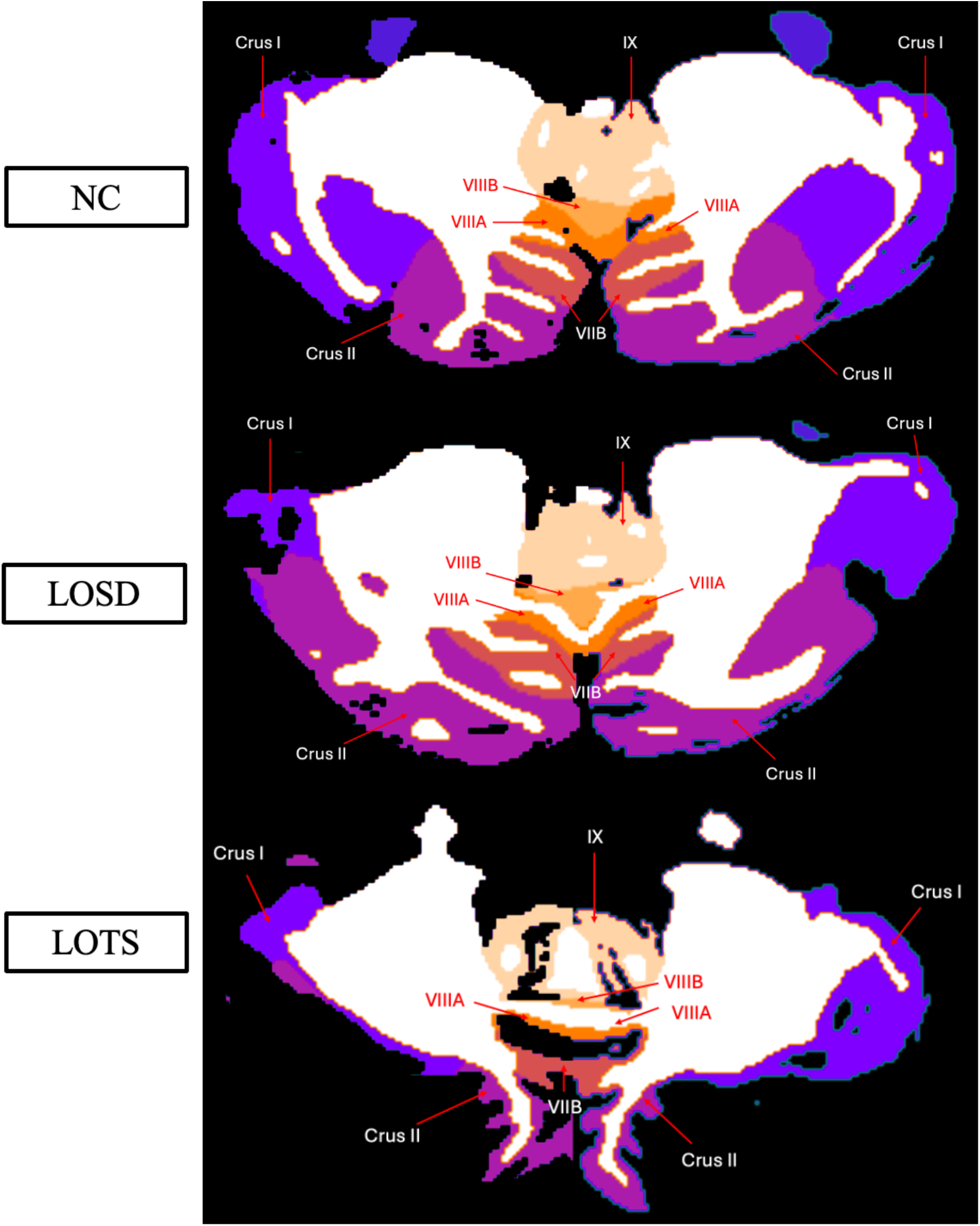
Labeled cerebellar lobule deep learning segmentation. Row one represents imaging from one 41- to 45-year-old neurotypical control (NC) female from the NIMH volunteer dataset. Row two represents imaging from one 41- to 45-year-old late-onset Sandhoff disease (LOSD) female patient. Row three represents imaging from one 41- to 45-year-old late-onset Tay-Sachs (LOTS) disease female patient. Each participants’ MRI scan was registered to the MNI space, and an axial slice at coordinate z = -40.5 was shown for each participant. Specific ages were redacted per Medrxiv requirements

### Supplement E: Cerebellar Lobule Volumetric Analysis

**Figure E1.**
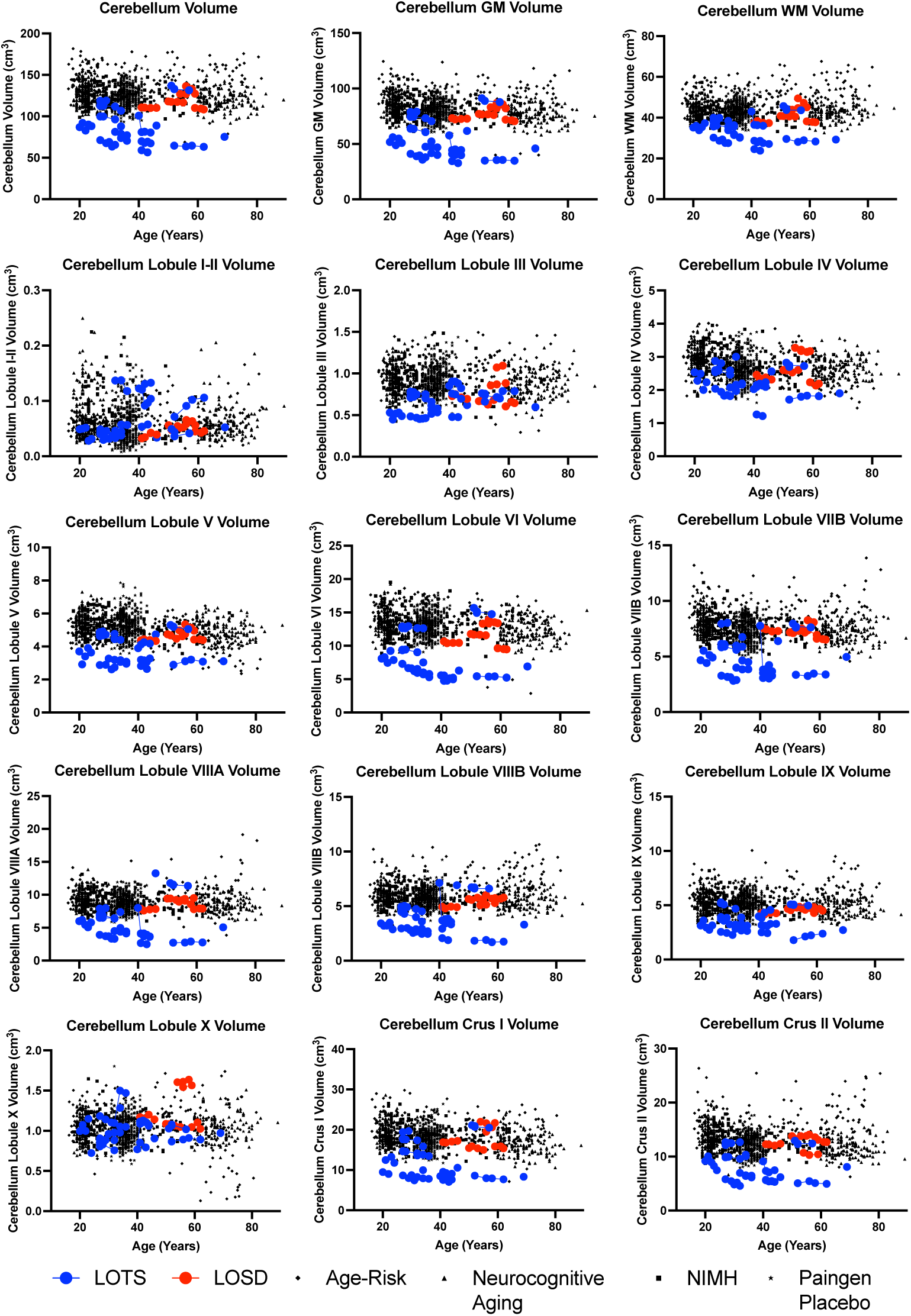
Volumetric Analysis of cerebellar lobule segmentation. Neurotypical controls are shown as individual black points with different symbol shapes for each Open-Source data set. Late-onset Sandhoff Disease (LOSD) patients are shown as red circles with red connecting lines. Late-onset Tay-Sachs (LOTS) disease patients are shown as blue circles with blue connecting lines.

### Supplement F: Cerebellar Lobule Cortical Thickness Analysis

**Figure F1.**
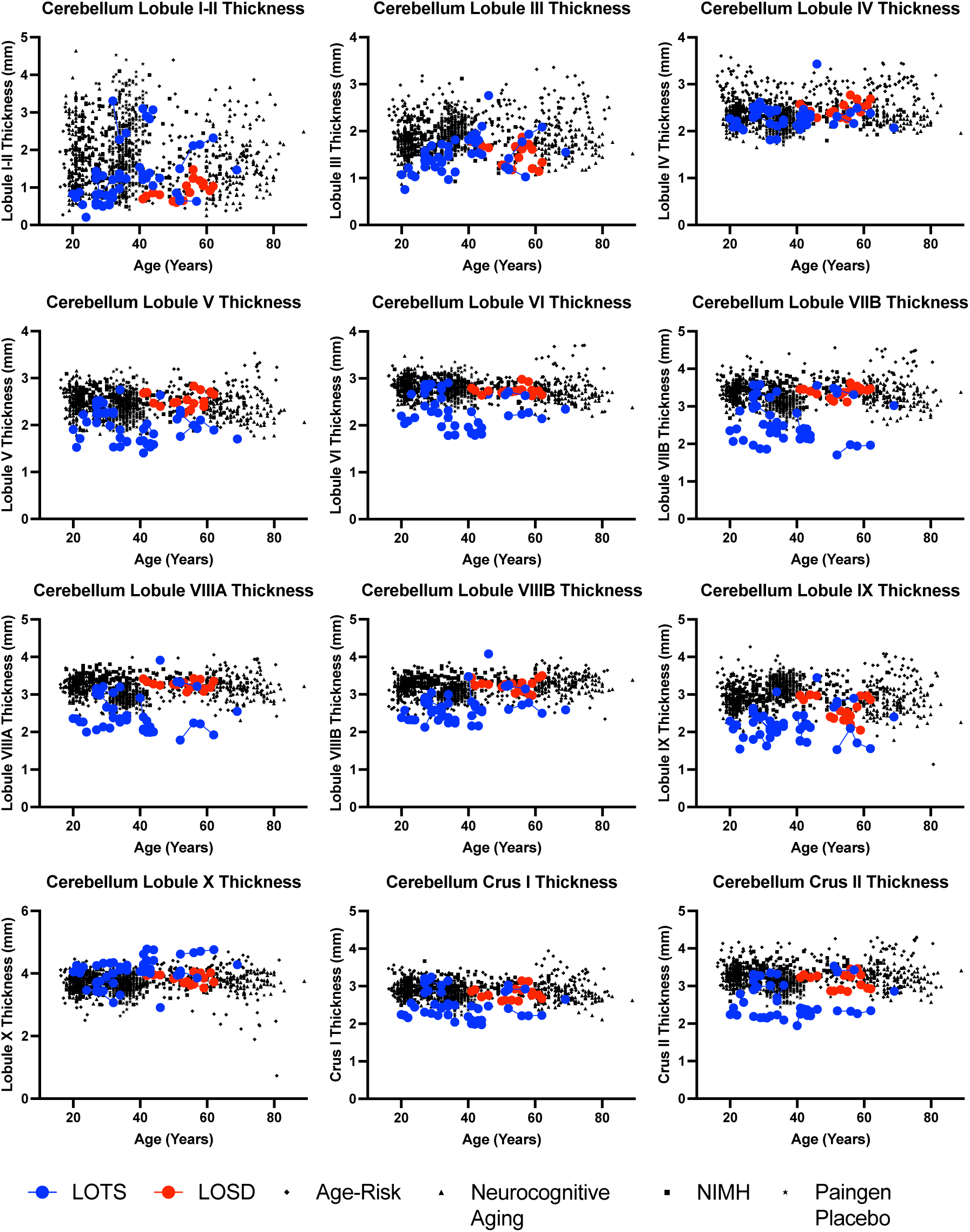
Cortical thickness analysis of the 12 cerebellar lobules. Neurotypical controls are shown as individual black points with different symbol shapes for each Open-Source data set. Late-onset Sandhoff Disease (LOSD) patients are shown as red circles with red connecting lines. Late-onset Tay-Sachs (LOTS) disease patients are shown as blue circles with blue connecting lines.

### Supplement G: Cerebellar Lobule Volumetric Analysis Estimates and Standard Errors

**Table G1.**
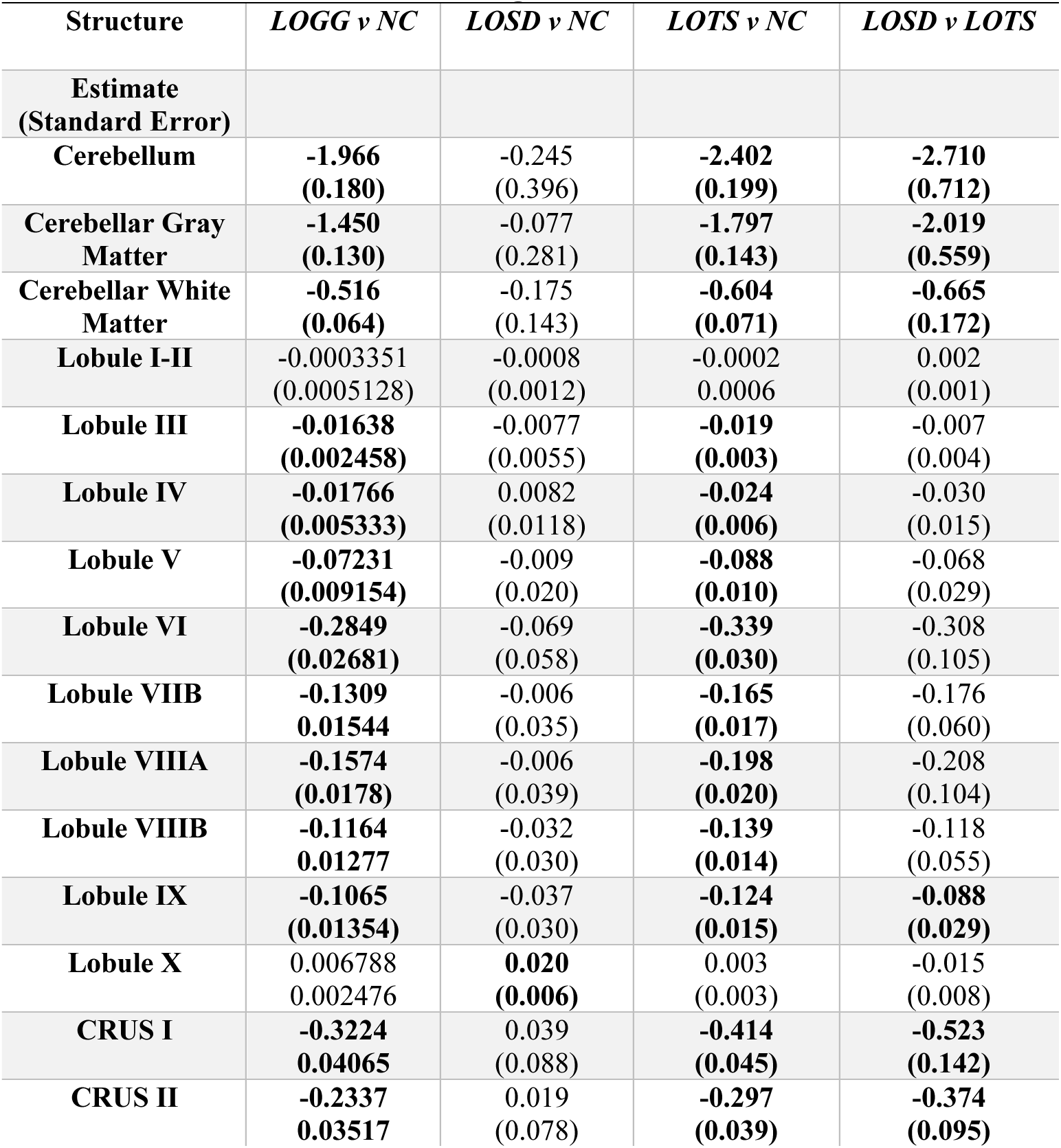
Cerebellar Volumetric MRI Analysis. All volumes were controlled for ICV. Statistical analysis was performed using a linear mixed effects model where *P*-values < 0.01 were considered significant and bolded.

### Supplement H: Cerebellar Lobule Cortical Thickness Analysis Estimates and Standard Errors

**Table H1.**
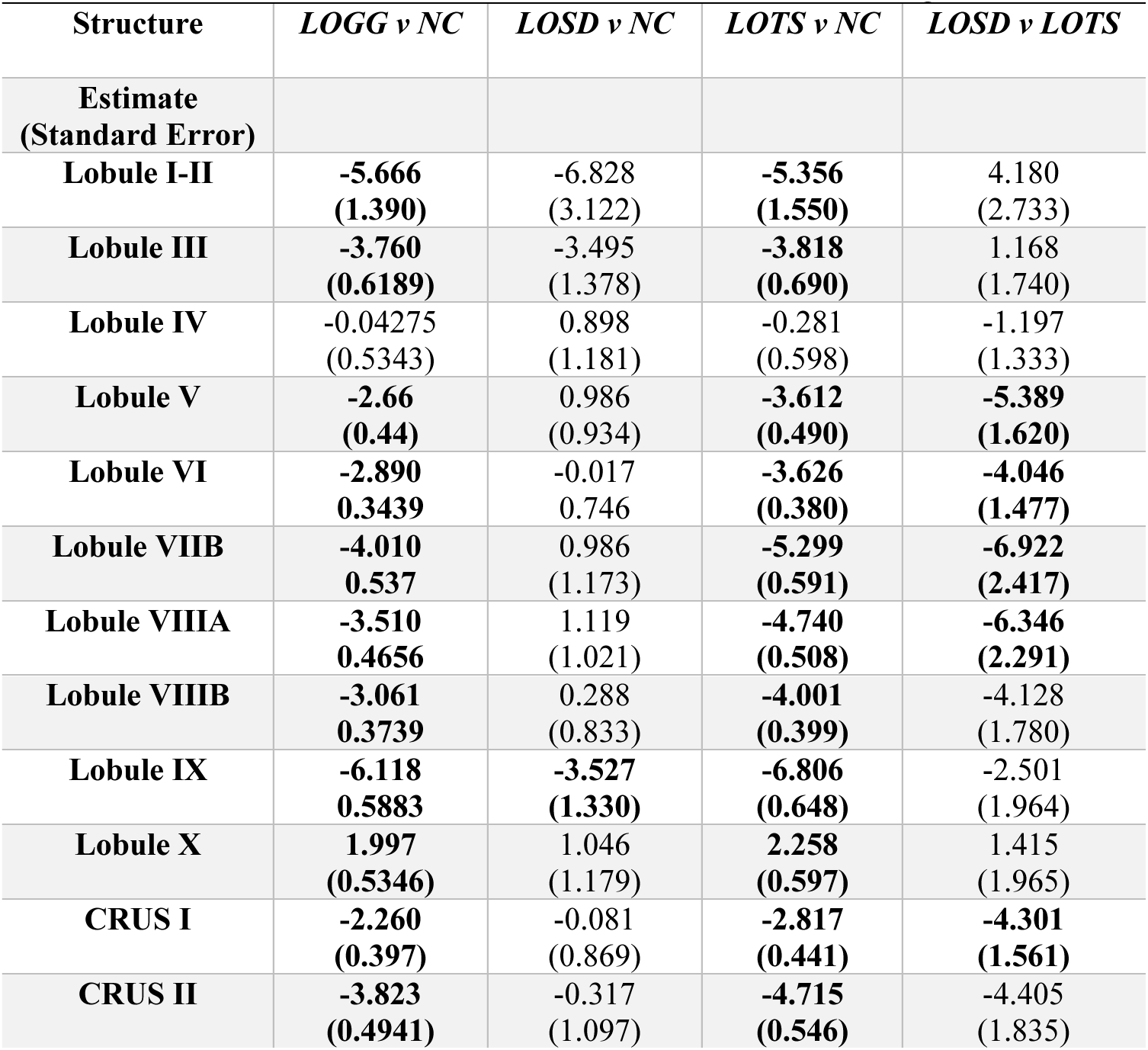
Cerebellar Cortical Thickness MRI Analysis. All volumes were all normalized in relation to the cube root of ICV (adimensional). Statistical analysis was performed using a linear mixed effects model where *P*-values < 0.01 were considered significant and bolded.

